# Mobile wastewater and environmental surveillance in low-resource settings: validating an integrated laboratory workflow for pathogen detection

**DOI:** 10.64898/2026.04.01.26349919

**Authors:** Andrea Bagi, Ananda Tiwari, Chikwendu Chukwudiegwu Mbachu, Dylan Shea, Tam T. Tran, Marc Christian Tahita, Palpouguini Lompo, Peter Mkama, Eric Lyimo, Vito Baraka, Alan Le Tressoler, Tarja Pitkänen, Adriana Krolicka

## Abstract

Mobile laboratories (MLs) offer a flexible platform for on-site wastewater and environmental surveillance, particularly in regions with limited laboratory capacity. Effective ML deployment, however, requires optimized molecular workflows that match local infrastructure, equipment, and workforce constraints. Here, we present an integrated ML workflow that combines Oxford Nanopore Technologies (ONT) sequencing for shotgun metagenomics, multiplex metabarcoding for microbial community profiling, and Biomeme-based qPCR for targeted pathogen detection. We introduce and validate a multiplexed ONT small subunit rRNA amplicon sequencing approach that enables simultaneous profiling of bacteria, archaea, and microeukaryotes in complex environmental samples. Sample pretreatment and nucleic acid extraction was tested for combined chemical–mechanical lysis followed by magnetic bead purification. Workflow performance was assessed by analyzing a known mock community (ZymoBIOMICS) and wastewater samples spiked with inactivated monkeypox virus (MPXV), demonstrating accurate taxonomic representation and sensitive MPXV detection. The multiplex metabarcoding workflow enabled rapid, high-resolution community profiling and showed stronger concordance with qPCR-based pathogen detection than whole-genome and whole-transcriptome amplification workflows. MPXV tiled amplicon sequencing was successfully incorporated into the ML protocol, supporting near-real-time genomic characterization in field settings. Across diverse environmental matrices, a multiple displacement amplification (MDA)-based metagenomic workflow combined with ML nucleic acid extraction reliably reproduced the composition of the mock community. Overall, integrating ONT sequencing, MDA-based metagenomics, multiplex metabarcoding, and mobile qPCR provides an efficient surveillance platform within a One Health framework. While key components of the workflow were validated under controlled laboratory conditions, pilot testing on environmental samples from sub-Saharan Africa demonstrates its applicability to real-world matrices.

**Graphical abstract:** 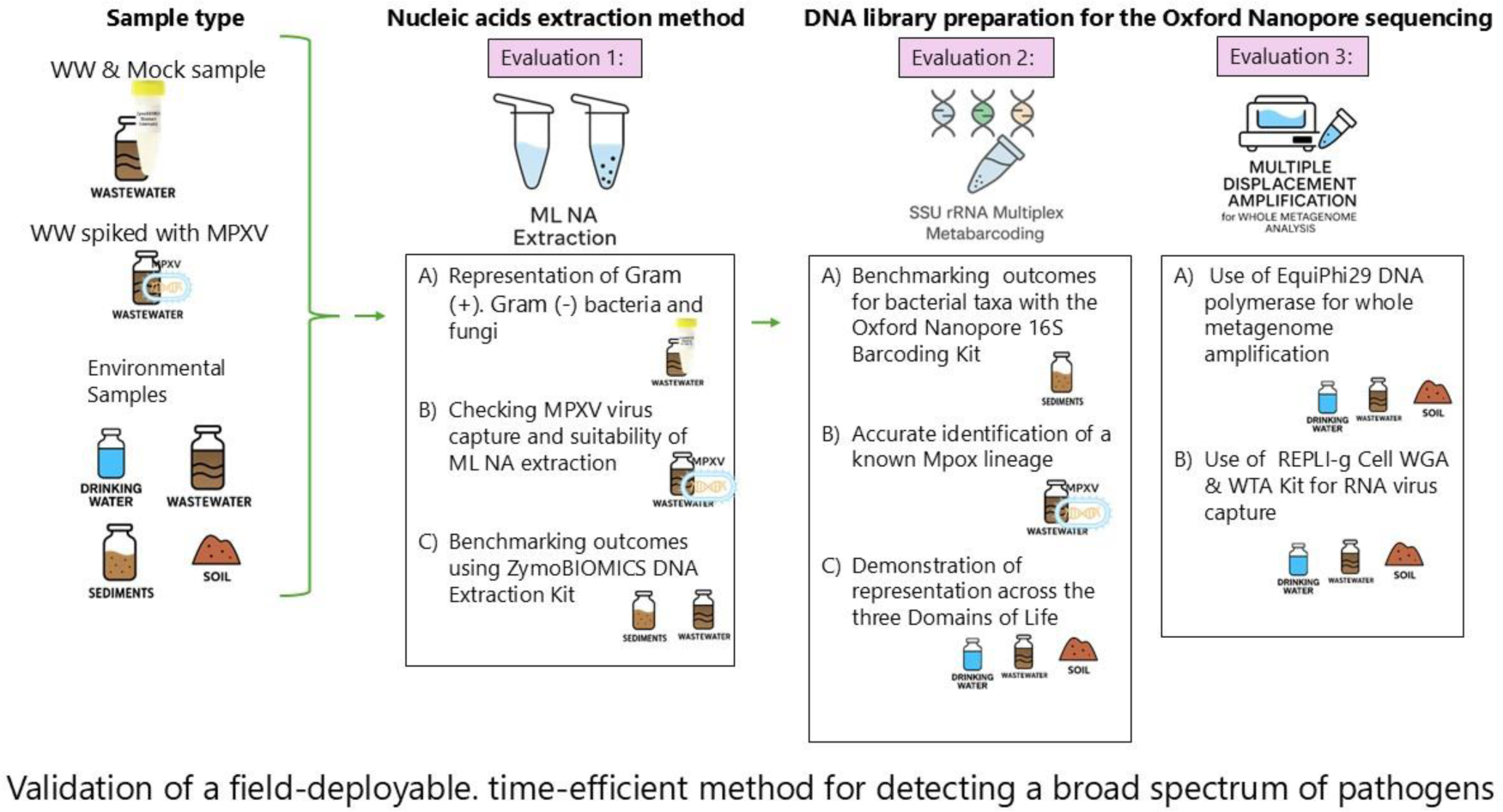

## 1 Introduction

Wastewater and environmental surveillance (WES) provides a powerful population-level approach for pathogen monitoring that complements classical clinical surveillance, particularly in resource-limited settings such as sub-Saharan Africa (SSA), where routine clinical systems may be limited or overextended^1–3^. However, implementing WES in SSA faces several challenges, including limited laboratory infrastructure, constrained availability of consumables, difficulties in cold-chain sample transport, and shortage of trained personnel, all of which can limit the timeliness of molecular surveillance^4–6^. Outbreaks in remote areas can further complicate centralized sampling strategies^7,8^.

Mobile laboratories (MLs), whether vehicle-mounted or rapidly deployable, offer practical solutions for on-site molecular analyses^6,8–10^. Successful ML deployment requires adapted and validated workflows that encompass sample pretreatment, nucleic acid (NA) extraction, and downstream analyses such as metabarcoding, metagenomics, and quantitative polymerase chain reaction (qPCR)^11^, followed by offline data analysis. Obtaining consistent microbial profiles across diverse environmental matrices depends on robust and validated sample-processing workflows. Effective NA extraction is critical for the accuracy and sensitivity of downstream qPCR and sequencing^12^. Extraction efficiency is influenced by factors such as the target NA (DNA/RNA), the sample matrix, processing speed, throughput, cost, and available equipment^11,13,14^.

Portable sequencing devices such as MinION and Flongle, combined with rapid barcoding kits, are well suited for field diagnostics^15,16^. However, low-biomass environmental samples (e.g., drinking water) pose challenges for library preparation, and often require additional steps to enrich environmental DNA^16,17^. Metabarcoding provides a cost-efficient means to profile bacterial, archaeal, and eukaryotic communities, but its applicability to viruses is limited due to the absence of universal viral marker genes^18–20^. Metagenomics enables comprehensive detection of bacteria, viruses, fungi, parasites, and antimicrobial resistance (AMR) genes and supports the identification of novel pathogens, although it remains relatively costly and bioinformatically demanding^21–24^. Quantitative PCR offers rapid and sensitive detection and quantification of targeted pathogens^22,25^, and portable systems such as the Franklin™ Biomeme qPCR platform enable field-deployable testing without reliance on cold-chain logistics^26,27^.

Over the past decade, several initiatives have supported ML networks in SSA^8,9,28,29^. For example, the East African Community ML network, established between 2017 and 2020 (https://www.eac.int/the-project/key-information), serves as a model system comprising mobile biosafety level (BSL)-3 and BSL-4 laboratories embedded within, and operated by, National Public Health Institutes^29^.

This study, conducted as part of the EU-funded ODIN and ODIN-MPOX projects^1^ (https://odin-wsp.tghn.org/), aimed to reposition mobile laboratories as decentralized platforms for environmental pathogen surveillance rather than standalone diagnostic units in SSA. The workflow integrates a portable ONT sequencing platform for shotgun metagenomics and multiplex metabarcoding, alongside a portable qPCR system (Biomeme). We evaluated a robust and reproducible analytical pipeline suitable for diverse wastewater and environmental matrices, including samples collected in Tanzania and Burkina Faso. Laboratory-based validation demonstrates the technical feasibility of this approach within a One Health framework and represents a critical step toward the practical integration of WES in resource-limited settings.

## 2 Materials and Methods

Portable analytical platforms and protocols organized in a modular design (Figure S1) formed the core of our ML workflow, including the MinION sequencer (Oxford Nanopore Technologies, ONT, UK) for metabarcoding and shotgun metagenomics, and the Biomeme mobile qPCR platform (USA) for targeted pathogen detection. Validation covered the full pipeline—NA extraction, ONT library preparation, metabarcoding, and qPCR—across diverse matrices to ensure broad applicability, including untreated, semi-treated, and treated wastewater, drinking water, soil, and sediment (Table S1). Detailed sampling and preprocessing are provided in the respective validation sections.

### 2.1 Validation of the nucleic acid extraction method

The ML NA extraction method was validated for: (1) recovery of high-quality NA with taxonomic coverage comparable to the commercial ZymoBIOMICS DNA/RNA Mini Kit (Zymo Research, USA); (2) accurate detection of Gram-positive and Gram-negative bacteria and fungi using the ZymoBIOMICS Microbial Community Standard; and (3) efficient recovery of MPXV, a key target pathogen in the ODIN-Mpox project. Detailed methods are provided in the subsections below.

#### 2.1.1 Validation of mobile lab DNA extraction against ZymoBIOMICS DNA Mini Kit

To compare NA yield, quality and taxonomic coverage of the ML NA extraction method to a commercially available ZymoBIOMICS DNA mini kit, wastewater and sediment samples were processed in parallel using both extraction protocols.

##### Sample collection

A 24 h composite influent wastewater sample was collected in May 2023 at the Mekjarvik wastewater treatment plant (WWTP) (Table S1) and stored at −80 °C. After thawing at 4 °C, 25 mL aliquots were filtered in triplicate through 0.22 µm nitrocellulose membranes and membranes were aseptically transferred to sterile tubes for subsequent NA extraction. A marine sediment sample collected in July 2021 near Kvitsøy (Norway) (Table S1) using a Van Veen grab was also included. Bulk surface sediment (0–2 cm) was collected from the smaller compartment using a sterilized spatula into a 50 mL tube and stored at -80°C. Prior to extraction, the sediment was thawed at 4°C, homogenized, and subsampled (1 g wet weight for ML; 0.5 g for ZymoBIOMICS). Sediment samples and wastewater filters were processed in parallel using both ML and ZymoBIOMICS NA extraction protocols, as described below.

##### Nucleic acid extraction – ML extraction method

The ML NA extraction method integrates chemical lysis (RLT buffer + DTT), mechanical disruption (needle homogenization), debris removal (syringe filtration), and magnetic bead–based purification (HighPrep PCR-DX MagBeads), followed by wash and elution steps. Briefly, lysis was performed by adding 1000 µL RLT Buffer (Qiagen, Germany) and 20 µL 2 M DTT (Fisher Scientific, USA) to a 5 mL tube containing filters and vortexing for ∼30 s. Lysates were transferred to a clean 2 mL tube and homogenized by aspirating and dispensing ∼10 times with a 1 mL syringe fitted with a 20G needle (BD, USA) followed by debris removal using a Millex-GV syringe filter (Millipore, USA) into a clean 2 mL tube. Nucleic acids were captured using HighPrep PCR-DX MagBeads (MagBio, USA) at a 1:1 bead-to-sample ratio, washed twice with RPE buffer (Qiagen, Germany), and eluted in 70 µL pre-warmed RNase-free water. A second purification step was performed by adding 70 µL isopropanol and 140 µL MagBeads, followed by RPE buffer (Qiagen, Germany) and 80% ethanol washes and elution in RNase-free water. Purified NAs were stored at −20 °C until further use.

##### ZymoBIOMICS kit extraction

For comparison with ML NA extraction method, DNA was extracted using the ZymoBIOMICS DNA Miniprep Kit (Zymo Research, USA) according to the manufacturer’s instructions. Extracted NAs were stored at −20 °C until further analysis.

##### Quantification and Quality Assessment

NA concentrations were measured using the Qubit™ dsDNA High Sensitivity Assay Kit (Thermo Fisher Scientific, USA). Purity was assessed with a NanoDrop™ One-C spectrophotometer, and integrity was evaluated by electrophoresis on 1.5–2% agarose gels. ***Library Preparation and Sequencing:*** Microbial composition in marine sediment samples was assessed by (1) full-length 16S rRNA gene amplicon sequencing and (2) a multiplex PCR-based metabarcoding approach. ***Full-length 16S rRNA metabarcoding*** libraries were prepared using the ONT 16S Barcoding Kit 24 V14 with ∼20 ng DNA as input. PCR reactions (50 µL) contained 10 µL barcode primer, 25 µL 2× KAPA3G Plant PCR Buffer, 0.4 µL KAPA3G Plant DNA Polymerase, and nuclease-free water. Cycling conditions were: 95 °C for 3 min; 25 cycles of 95 °C for 20 s, 56 °C for 40 s, and 72 °C for 1 min 40 s; followed by a final extension at 72 °C for 10 min. Amplicons were purified, quantified, pooled equimolarly, and sequenced on a MinION Mk1B using a Flongle flow cell (FLO-MIN114). M***ultiplex PCR-based metabarcoding*** libraries were prepared using primers listed in Table S2 together with the Native Barcoding Kit (ONT, UK). Each 25 µL reaction contained 2 µL DNA, 800 nM of each primer, 12.5 µL 2X KAPA Plant PCR Buffer, 1 U KAPA3G Plant DNA Polymerase, and nuclease-free water. Cycling conditions matched those used for full-length 16S library preparation. Amplicons were purified with AMPure XP beads, eluted in 20 µL EB, then barcoded and prepared for sequencing following the Native Barcoding Kit protocol. Adapter-ligated amplicons concentrations were quantified using a NanoDrop One-C before pooling. Libraries were loaded onto a Flongle flow cell (FLO-MIN114) and sequenced on a MinION device (ONT, UK). Sequencing runs for both full-length ONT-16S and Multiplex PCR libraries were performed in MinKNOW v24.02.8, with a minimum read length of 200 bp, Q score ≥ 10, and super-accurate basecalling (400 bps). Wastewater extracts were not sequenced; only DNA yield and quality metrics were evaluated.

##### Bioinformatic processing

Raw sequencing reads from (1) full-length 16S rRNA gene libraries and (2) multiplex PCR libraries were processed using the epi2me-labs/wf-16s workflow executed through the EPI2ME Labs interface (version 24.08-01). Reads shorter than 800 bp were removed prior to analysis. Taxonomic assignment was performed using minimap2 (version 2.26-r1175) against the SILVA database (SILVA release 138.1) as implemented in the workflow^30^.

#### 2.1.2 Mock community-based validation (via MDA and ONT metagenomics)

##### Sample collection and processing

The efficacy of the ML-NA extraction method in capturing diverse microbial communities within complex environmental matrices was evaluated using the ZymoBIOMICS Microbial Community Standard, which includes inactivated Gram-positive and Gram-negative bacteria and fungi (Table 1). DNA was extracted from 24h composite wastewater samples (after prior filtration through 0.22 µm nitrocellulose membranes, Whatman™ 25 mm) and the ZymoBIOMICS Microbial Community Standard (Zymo Research, USA) using the ML-NA protocol (Section 2.1.1). Triplicate wastewater extractions were pooled, and DNA concentrations were normalized to 5 ng/µL before spiking wastewater DNA with mock community DNA at 1:10 and 1:100 (v/v) ratios.

**Table 1.**
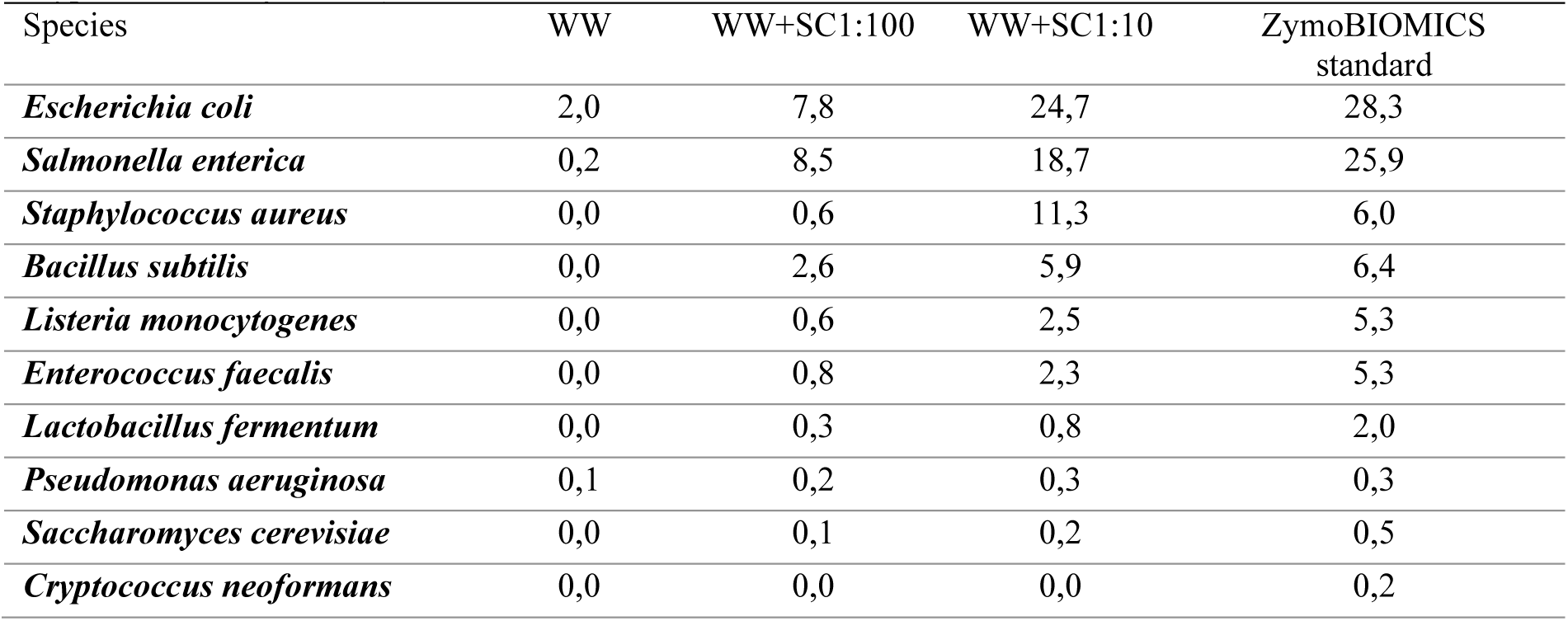
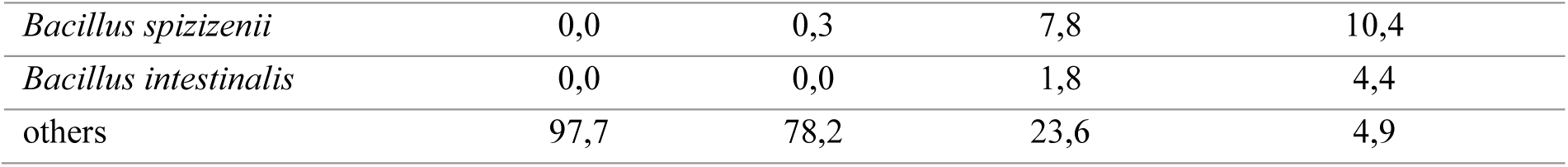
Relative abundance (%) of microbial species in sequencing reads from the ZymoBIOMICS standard and wastewater samples. WW: wastewater; wastewater spiked with ZymoBIOMICS at 1:100; WW-SC1:10; and ZymoBIOMICS standard. Species present in the ZymoBIOMICS standard in bold font *(Listeria monocytogenes, Pseudomonas aeruginosa, Bacillus subtilis, Escherichia coli, Salmonella enterica, Lactobacillus fermentum, Enterococcus faecalis, Staphylococcus aureus, Saccharomyces cerevisiae, Cryptococcus neoformans*)

##### Library Preparation and Sequencing

DNA was enriched by multiple displacement amplification (MDA) using high-fidelity EquiPhi29 DNA Polymerase and 5′-phosphorylated random hexamers (ThermoFisher, USA) following the manufacturer’s protocol. MDA products were purified with HighPrep PCR-DX MagBeads (MagBio, USA) at a 1:1 bead-to-sample ratio, washed twice with 80% ethanol, and eluted in 60 µL H₂O. Libraries were prepared using the Rapid Barcoding Kit (ONT, UK) and sequenced on a MinION device with an R10.4.1 flow cell. Raw reads were basecalled and quality-filtered (minimum length 200 bp) in high-accuracy mode using MinKNOW v23.07.15.

##### Bioinformatic processing

Raw reads were processed using the wf-metagenomics workflow with default settings in EPI2ME Labs (version 24.08-01) with taxonomic assignments generated using the PlusPF-8 database at the species level^30–32^ (https://github.com/nanoporetech/medaka; https://github.com/pysam-developers/pysam)

#### 2.1.3 Validation of MPXV capture and accurate phylogenetic identification

To evaluate the efficiency of the ML-NA extraction method, paired experiments were performed in which spiked and unspiked wastewater samples were processed using the ML-NA extraction protocol. Inactivated Mpox virus (MPXV, clade II, (https://www.european-virus-archive.com/evag-portal/ictv-taxonomy/monkeypox-virus) served as the spiking material. Two viral loads were tested, by adding 100 µL or 200 µL of virus stock, estimated at 10⁶ TCID₅₀/mL prior to inactivation.

##### Sample collection and processing

The wastewater influent sample used in this experiment was collected in April 2025 from the Breivika WWTP in Tromsø, Norway (Table S1). Prior to NA extraction, the wastewater sample was filtered through a sterile syringe fitted with a 0.22 µm Whatman™ 25 mm nitrocellulose membrane filter. Approximately 25 mL of wastewater was passed through the filter under gentle pressure to minimize cell lysis. Membranes were then transferred aseptically to sterile tubes for NA extraction. Known quantities of inactivated MPXV were either (1) spiked into the wastewater sample prior to filtration or (2) applied directly onto the filter after filtration. The unspiked wastewater sample served as a negative control.

##### Endpoint ARTIC PCR

The presence of MPXV DNA was assessed by PCR amplification followed by electrophoresis on 0.8% agarose gels. Viral DNA was amplified using an ARTIC-style tiled PCR approach to obtain near-complete genome coverage. Two primer pools, each containing ∼24 primers, generated overlapping amplicons across the MPXV genome (∼12 amplicons of ∼8.5 kb per pool) (Table S3). PCR reactions (25 µL) were prepared using LongAmp™ Hot Start Taq 2X Master Mix (New England Biolabs, USA), with appropriate positive and negative controls. Thermocycling conditions were: 94 °C for 30 s; 25 cycles of 94 °C for 30 s, 55 °C for 40 s, and 65 °C for 9 min; followed by a final extension at 65 °C for 10 min. Amplicon integrity was verified by agarose gel electrophoresis, and samples showing nonspecific amplification or contamination were excluded.

##### Amplicon Cleanup, Library preparation, Sequencing and Bioinformatic processing

PCR products from both primer pools were combined per sample and purified using HighPrep PCR-DX MagBeads (MagBio, USA) at a 1:1 bead-to-sample ratio. Purified DNA was eluted in 15 µL EB buffer (Qiagen, Germany). Sequencing libraries were prepared using the Rapid Barcoding Kit (ONT, UK) and sequenced on a MinION device with an R10.4.1 flow cell. Raw reads were basecalled and filtered in high-accuracy mode using MinKNOW v23.07.15 with a minimum read length of 200 bp^33^. Sequencing data were analyzed using two publicly available bioinformatic tools: the artic-mpxv-nf workflow for generating consensus MPXV genomes from pooled tiled amplicons (https://github.com/artic-network/artic-mpxv-nf), and Squirrel for sequence alignment and maximum-likelihood phylogenetic inference^31,32,34,35^.

### 2.2 Validation of Library Preparation for Microbial Community Characterization

The samples used for NA extraction validation in Section 2.1.1 (Table S1) were also used to validate microbial characterization using multiplex metabarcoding with high-throughput amplicon sequencing, and shotgun metagenomics. For multiplex metabarcoding, library preparation was evaluated using primers targeting the small-subunit (SSU) rRNA genes of bacteria^36^, archaea^37^, and eukaryotes^38^. Expected amplicon sizes were ∼1,465 bp for bacteria, ∼1,492 bp for archaea, and ∼1,542 bp for eukaryotes (Table S2).

#### 2.2.1 Benchmarking multiplex metabarcoding against ONT-16S Kit

To validate the Multiplex PCR metabarcoding approach, we compared taxonomy coverage between two library preparation methods: (1) multiplex metabarcoding and (2) the commercially available 16S Barcoding Kit 24 V14 (ONT, UK). Sample collection, library preparation, sequencing, and bioinformatic procedures are described in Section 2.2.1. Both ML-NA and ZymoBIOMICS DNA Miniprep kit extracts from sediment were included.

#### 2.2.2 Benchmarking the multiplex metabarcoding against ONT metagenomics

##### Sample collection and processing

Environmental samples, including drinking water, untreated (WW-U), semi-treated (WW-S), treated wastewater (WW-T), and soil were collected in June 2024 from active surveillance sites near WWTPs in the Mabibo, Masani, and Vingunguti wards of Dar es Salaam, Tanzania) (Table S1). Drinking water (750–3000 mL; drinking water; DW) and wastewater (10–30 mL; WW) samples were filtered in triplicate through 0.22 µm polyethersulfone Sterivex-GP cartridges (Millipore, USA). Drinking water samples were filtered using a peristaltic pump (Watson Marlow, UK), and wastewater samples were filtered with sterile 50 mL syringes. Filters were aseptically removed, fragmented, and transferred to 2 mL tubes for NA extraction. Triplicate ∼5 mL surface soil samples were also collected for NA extraction. NA extraction followed the ML-NA protocol described in Section 2.1.1.

##### Library preparation, sequencing

For multiplex metabarcoding, triplicate NA extracts were pooled and amplified using primers targeting the SSU rRNA genes: 16S rRNA for bacteria and archaea and 18S rRNA for eukaryotes (Table S2). PCR reactions contained 2 µL DNA template, 800 nM of each primer, 1× HS LongAmp Taq polymerase, and nuclease-free water. Cycling conditions were 95 °C for 30 s; 32 cycles of 95 °C for 20 s, 56 °C for 40 s, and 65 °C for 1 min 40 s; followed by a final extension at 65 °C for 10 min. Amplicons were quantified using the Qubit™ HS DNA Assay Kit (Thermo Fisher Scientific, USA) and checked against a negative PCR control. Biological triplicates were pooled and purified in a 96-well format using HighPrep PCR-DX magnetic beads (MagBio, USA), washed twice with 80% ethanol, and eluted in 40 µL nuclease-free water. Sequencing libraries were prepared using the Rapid Barcoding Kit (ONT, UK), then sequenced on a MinION device using an R10.4.1 flow cell. Raw reads were basecalled and filtered (minimum length 200 bp) in high-accuracy mode using MinKNOW v23.07.15.

#### 2.2.3 EquiPhi-based multiple displacement amplification and shotgun metagenomics

Environmental samples were collected in September 2024 from an active pathogen surveillance site in Ouagadougou, Burkina Faso and used for testing of MDA enrichment and shotgun metagenomics (Table S1). Surface water (750–1000 mL), drinking water (2500 mL), and wastewater (25 mL) samples were filtered in triplicate through 0.22 µm Sterivex-GP cartridges (polyethersulfone; Millipore). Surface water and drinking water samples were filtered using a peristaltic pump as described in section 2.2.2. Soil samples were also collected for NA extraction as described in section 2.2.2. DNA was extracted from soil samples and filters using the ML-NA extraction protocol (see Section 2.1.1), and individual NA extracts were pooled in preparation for MDA and shotgun metagenomic sequencing.

##### Library preparation and sequencing

Total NA from environmental samples underwent whole-metagenome amplification (WMA) using the EquiPhi Phi29 DNA Polymerase Kit (Thermo Fisher Scientific, USA). Samples were denatured and primed with exonuclease-resistant random hexamers before isothermal amplification according to the manufacturer’s protocols. DNA quality before and after amplification was assessed using a Qubit fluorometer with the dsDNA HS kit fluorometric quantification (Thermo Fisher Scientific, USA). Amplified DNA was purified with HighPrep PCR magnetic beads (MagBio, USA), treated with T7 Endonuclease I (New England Biolabs, USA) to linearize branched and hyper-branched Phi29 amplification products, and subjected to a second bead cleanup to obtain high-quality linear DNA. Nanopore sequencing libraries were prepared using the ONT Native Barcoding Kit with end-repair and dA-tailing, barcode ligation, equimolar pooling, and adapter ligation, with magnetic-bead cleanups between steps. Pre-sequencing quality control included quantification of adapter-ligated DNA, evaluation of barcode balance, and checking fragment-size distributions. Libraries were loaded onto ONT flow cells and sequenced on a MinION device using MinKNOW. Run-level quality control monitored pore activity, read-length profiles, and Q-scores. Basecalling and demultiplexing followed ONT’s standard pipeline and only reads meeting recommended quality thresholds were retained.

##### Bioinformatic processing

Metagenomic data analysis was conducted using the EPI2ME Labs metagenomic workflow, with antimicrobial resistance gene (ARG) mapping enabled and annotation performed using the AMRFinder reference database.

#### 2.2.4 REPLI-g Cell Whole Genome Amplification & WTA workflow

Environmental samples from Dar es Salaam (Table S1, wastewater: 10M-12M, drinking water: 1M) were extracted using the ML-NA extraction protocol and subsequently enriched with the REPLI-g Cell WGA & WTA Kit (Qiagen, Germany) to enable parallel whole-genome amplification (WGA) and whole-transcriptome amplification (WTA). NA aliquots were split into WGA and WTA reactions following the manufacturer’s protocol. Genomic DNA was amplified isothermally via MDA, and RNA underwent reverse transcription prior to WTA. After amplification of DNA and ligase-treated cDNA amplification, NA yield and purity were assessed using a Nanodrop spectrophotometer (Thermo Scientific, USA). Amplified products were purified with HighPrep PCR magnetic beads (MagBio, USA) and treated with T7 endonuclease to remove hyper-branched structures. DNA concentration was measured using the Qubit High-Sensitivity (HS) DNA kit before library preparation. Sequencing libraries were prepared using the ONT Native Barcoding Kit following the manufacturer’s protocol. Sequencing and bioinformatic processing were performed as described in section 2.2.2.

### 2.3 Validation of Biomeme-based qPCR

Samples used for NA extraction validation (Section 2.1.1; Table S1) were also used to validate targeted, sensitive analysis using Biomeme-based qPCR for ML workflow.

#### 2.3.1 Selection, and integration of qPCR assays

Eight priority pathogens, identified through stakeholder surveys in the DRC, BF, and TZ, were targeted for validation using the ML qPCR workflow^1^. and were targeted using the ML qPCR workflow for validation. Due to its portability and minimal infrastructure needs, the Franklin Biomeme platform was chosen for the mobile qPCR workflow^26,27,39^. The instrument operates on internal or external power, reducing dependence on stable electricity^27,39^. Thermocycling is controlled via a smartphone app for real-time monitoring, direct data export, and optional cloud upload when internet is available^27,39^. Its nine-well format enables multiplex detection across green, amber, and red channels, supporting up to three targets per sample.

#### 2.3.2 Implementation of qPCR assays and PCR reactions optimalization

qPCR assays were selected based on published literature for the priority pathogens for ML (Table S4). To validate these assays, including targets for Mpox (clades Ia, Ib, II, and generic Mpox, Orthopoxvirus), *Klebsiella pneumoniae*, *Salmonella Typhi*, toxigenic *Vibrio cholerae*, *Escherichia coli*, poliovirus, rotavirus A, and hepatitis A virus, synthetic DNA fragments containing target sequences were designed and ordered (Table S5). All assays were standardized to uniform thermocycling conditions to enable multiplexing and maximizing the capacity of the Biomeme thermocycler. Pathogen qPCR assays were validated for ML use by optimizing master mix, primer–probe concentrations, annealing temperature, and assay efficiency. Initial optimization was conducted in a static laboratory using droplet digital PCR (ddPCR) to enable broader testing of conditions. To ensure comparability between mobile and static laboratory results, SsoAdvanced Universal Probes Supermix (Bio-Rad, USA) was used instead of the Biomeme LyoDNA mix. Assays were evaluated across annealing temperatures (50–63 °C) to identify a common, robust condition.

##### qPCR

Assay efficiency on the Biomeme instrument was evaluated using standard curves generated from 10-fold serial dilutions of synthetic DNA targets (Table S5). Reactions (20 µL) contained 10 µL SsoAdvanced Universal Probes SuperMix (Bio-Rad, USA), 400 nM primers, 200 nM probe, and 2 µL template DNA. All reactions followed a unified thermocycling program: 95 °C for 6 min, then 45 cycles of 95 °C for 5 s and 60 °C for 20 s. Six dilution points (∼10⁵–10⁰ gene copies/µL) and three no-template controls were included per run. Due to the 9-well format, standards were run as single replicates. Ct values were plotted against log₁₀ gene copy number, and linear regression was used to estimate slope and intercept. Amplification efficiency (E) was calculated from the slope: E = 10(−1/m) – 1. Equations derived from standard curves were subsequently used for calculating gene copy (GC) concentrations per assays per sample following established qPCR conventions ^40^.

#### 2.3.3 Pilot testing of field-collected samples

To evaluate assay performance across environmental matrices, optimized qPCR assays were applied to selected samples from Tanzania, including drinking water, wastewater, and soil (Table S1). DNA extracts were screened for target pathogens under reaction conditions outlined in 2.3.1 to assess the presence of target pathogens across matrices. Results were compared with pathogen signals from metagenomic shotgun sequencing for the same samples. Agreement between qPCR and sequencing was assessed by comparing the presence or absence of target agents across methods.

### 2.4 Statistical analysis and visualization

Statistical analyses were conducted in R (version 4.5.2) using the tidyverse packages for data handling and ggplot2 (version 2_4.0.1) for visualization. For sequencing data, relative abundances were calculated by normalizing read counts within each sample. Differences in taxonomical abundance across sample types were tested using a one-way ANOVA applied to arcsine-square-root-transformed relative abundances. Differences in community composition between NA extraction methods and between library preparation approaches were assessed using permutational multivariate analysis of variance (PERMANOVA) based on Bray–Curtis dissimilarities, implemented using the adonis2 function in the vegan package. Sequencing reads corresponding to specific pathogens were identified by matching species names and full taxonomy strings to a curated pathogen list using regular-expression rules. Venn diagrams were generated to visualize variation in pathogenic communities related to contrasting methodologies ^41^.

## 3 Results

### 3.1 Validation of the Nucleic Acid Extraction method

#### 3.1.1 Validation of mobile lab DNA extraction against ZymoBIOMICS DNA Mini Kit

Compared with the ZymoBIOMICS DNA Miniprep Kit, the ML workflow produced slightly higher-quality DNA from both wastewater and sediment samples, shown by clearer bands on agarose gels, while maintaining similar purity across sample types (Figure S2A–C). Nucleic acids extracted from wastewater showed mean concentrations of 14.1 ± 2.1 ng/µL (Qubit) with 260/280 and 260/230 ratios of 2.08 ± 0.10 and 0.93 ± 0.17, respectively, for the ZymoBIOMICS kit, compared with higher yields of 20.4 ± 3.8 ng/µL and improved purity (260/280 = 2.19 ± 0.02; 260/230 = 2.35 ± 0.06) obtained using the ML NA extraction method (Table S7). In sediment samples, DNA concentrations were higher with Zymo (25.7 ± 2.2 ng/µL) than with ML (12.8 ± 1.5 ng/µL), whereas purity was comparable across extraction methods (NanoDrop 260/280: Zymo 1.95 ± 0.02, ML 1.99 ± 0.02; 260/230: Zymo 1.54 ± 0.25, ML 1.41 ± 0.0). The ML workflow consistently yielded high-molecular-weight DNA with reduced degradation in both sample types, as confirmed by gel electrophoresis (Figure S2B), supporting its suitability for metagenomic library preparation and long-read sequencing.

##### Taxonomy coverage

Microbial communities were similar across extraction methods for both 16S and multiplex sequencing data. At the phylum and family levels, overall community composition was largely consistent between methods, with alpha and beta diversity analyses confirming comparable richness and structure (Figure 1A). Most bacterial taxa (67–68%) were shared between workflows (Figure 1B). Overall, the ML-NA extraction provided high-quality DNA with superior integrity for long-read sequencing, with taxonomic recovery broadly comparable to a standard commercial kit. LEfSe analysis (FDR-adjusted p < 0.05) identified taxa distinct to each method. (Figure S3).

**Figure 1.**
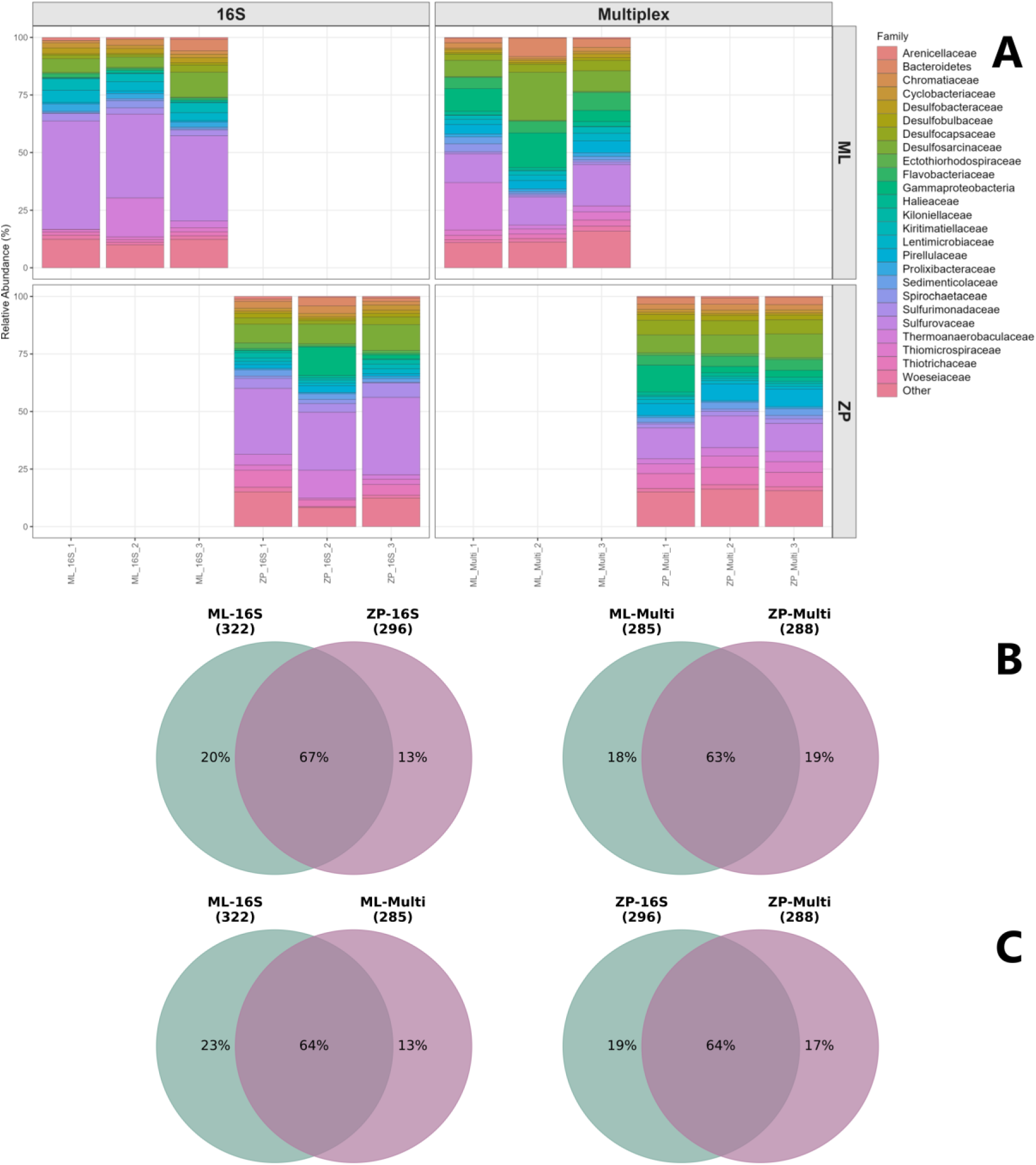
Benchmarking of Multiplex PCR metabarcoding and ONT-16S Barcoding kit. (**A**) Microbe community composition obtained from ONT-16S (left panels) and Multiplex PCR metabarcoding (right panels) sequencing of ML-extracted (ML) and ZymoBIOMICS-extracted (ZP) samples. (**B**) Venn diagrams depicting unique and shared bacterial taxa across extraction methods and sequencing approaches. Note: The diagram shows the percentage of bacterial features detected with >1 read in all three replicates for the two extraction methods: ML (mobile laboratory workflow) and ZP (ZymoBIOMICS kit). Left: Oxford Nanopore 16S kit; Right: library preparation using a multiplex primer set. (**C**) Venn diagrams of bacterial features shared between library preparation methods. Features with ≥1 read in all three replicates are shown for the 16S kit (Oxford Nanopore) and the Multiplex primer set. Left: Mobile laboratory (ML) protocol; Right: ZymoBIOMICS (ZP) protocol.

#### 3.1.2 Mock community-based validation (via MDA and ONT metagenomics)

Average DNA concentrations were 170 ng/µL for the ZymoBIOMICS standard and 89 ng/µL for ML workflow. Sequencing began with 620 active pores and generated ∼693.65 Mb across 268,220 reads (N50: 4.34 kb), yielding 8.47 GB of raw data. After basecalling (Q score ≥ 9), 543.07 Mb of reads passed quality filtering, while 66.36 Mb failed. On average, each sample produced ∼130 Mb of data and ∼68,000 reads, with 96% of reads passing quality control, indicating high sequencing performance and data quality.

In the mock community, *E. coli* was the most abundant taxon (28.3%). In wastewater extracts spiked with 10% mock community DNA, *E. coli* remained dominant (24.7%), whereas in samples spiked with 1%, it was the second most abundant taxon (7.8%). The relative contribution of native wastewater bacteria decreased from 97.7% (no spike) to 78.2% (1% spike) and 23.6% (10% spike), demonstrating accurate recovery of relative abundances.

Overall, the ML workflow reliably captured both community composition and relative abundance in wastewater while producing high-quality sequencing data suitable for downstream analyses. The *Bacillus subtilis* group includes closely related species with substantial genomic similarity, which limits species-level resolution in metagenomic classification. This is likely to explain the increased apparent diversity of *Bacillus* species observed in this dataset (Table 1).

#### 3.1.3 Validation of MPXV capture and accurate phylogenetic identification

Inactivated MPXV was consistently detected in all spiked wastewater samples across experimental setups, while non-spiked samples remained negative (Figure S2D), confirming efficient recovery of MPXV DNA by the ML extraction workflow. Post-sequencing analyses showed robust phylogenetic clustering of the generated Mpox sequences with both public reference genomes and the supplier-provided genome (Figure 2; Figure S4).

**Figure 2.**
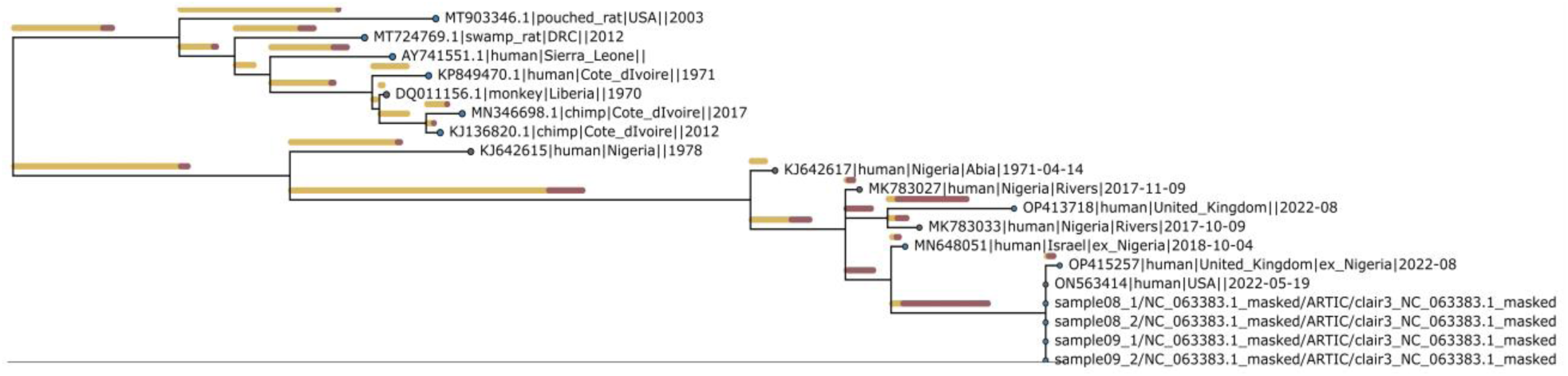
Phylogenetic alignment of Mpox-spiked wastewater replicates with reference sequences. Multiple Mpox-spiked wastewater replicates cluster with reference Mpox virus sequences generated using the Squirrel tool.

Accurate phylogenetic placement was achieved with as little as 4.5 MB of sequencing data, compared with ∼200 MB in standard runs. Both Flongle and MinION flow cells enabled reliable Mpox characterization (Figure S4). Bioinformatic analysis using either the EPI2ME Labs interface or command-line workflows produced consistent results, although software version differences affected graphical outputs. Likewise, the artic-mpxv-nf pipeline yielded identical phylogenetic results when run with either the default or custom primer scheme (Figure S4).

### 3.2 Validating library preparation and sequencing approaches

#### 3.2.1 Benchmarking multiplex metabarcoding against ONT-16S Kit

When benchmarking bacterial community profiling, read-length distributions differed between library preparation methods: the ONT 16S kit produced a sharp peak at ∼1.6 kb, while the multiplex approach showed a broader distribution centered at ∼1.47 kb, reflecting variable amplicon sizes. Despite this, both methods recovered highly similar community compositions at the phylum and family levels (Figure 1A).

Principal Coordinates Analysis (PCoA) based on Bray–Curtis dissimilarities showed tight clustering of technical replicates (PERMANOVA, R² = 0.95, p = 0.001), indicating high reproducibility. Pairwise PERMANOVA detected no significant differences at either the feature or family level (p > 0.05). In sediment samples, 64% of bacterial features were shared between the two library preparation approaches (Figure 1B).

Across six samples amplified with the same primer set, LEfSe identified several differentially abundant families (FDR-adjusted p < 0.05) (Figure S3). Overall, high-level taxonomic profiles were consistent across methods, with observed differences mainly reflecting relative abundance shifts rather than compositional disagreement.

#### 3.2.2 Benchmarking the multiplex metabarcoding against ONT metagenomics

For benchmarking, multiplex metabarcoding was compared with non-enriched ONT metagenomics. The multiplex sequencing run (samples M1–M15; Table S1) generated ∼1.59 Gb of data (2.13 million reads; N50 = 933 bp), of which 1,805,037 reads passed quality filtering. Approximately 17% (305,338 reads) remained unclassified. Samples yielded an average of 128,931 reads (range: 87,750–184,274), and all but sample M7 reached saturation, indicating sufficient sequencing depth.

Alpha diversity (Chao1) was significantly lower in drinking water (DW) compared with wastewater and soil samples (ANOVA: p = 0.0007; F = 13.804). Domain-level composition varied by sample type (Figure 3A). DW samples (M1–M3) showed relatively higher proportions of *Eukaryotes* and *Archaea*, including *Thaumarchaeota*, *Crenarchaeota*, and *Euryarchaeota,* groups known to persist in oligotrophic and engineered water systems^20^.

**Figure 3.**
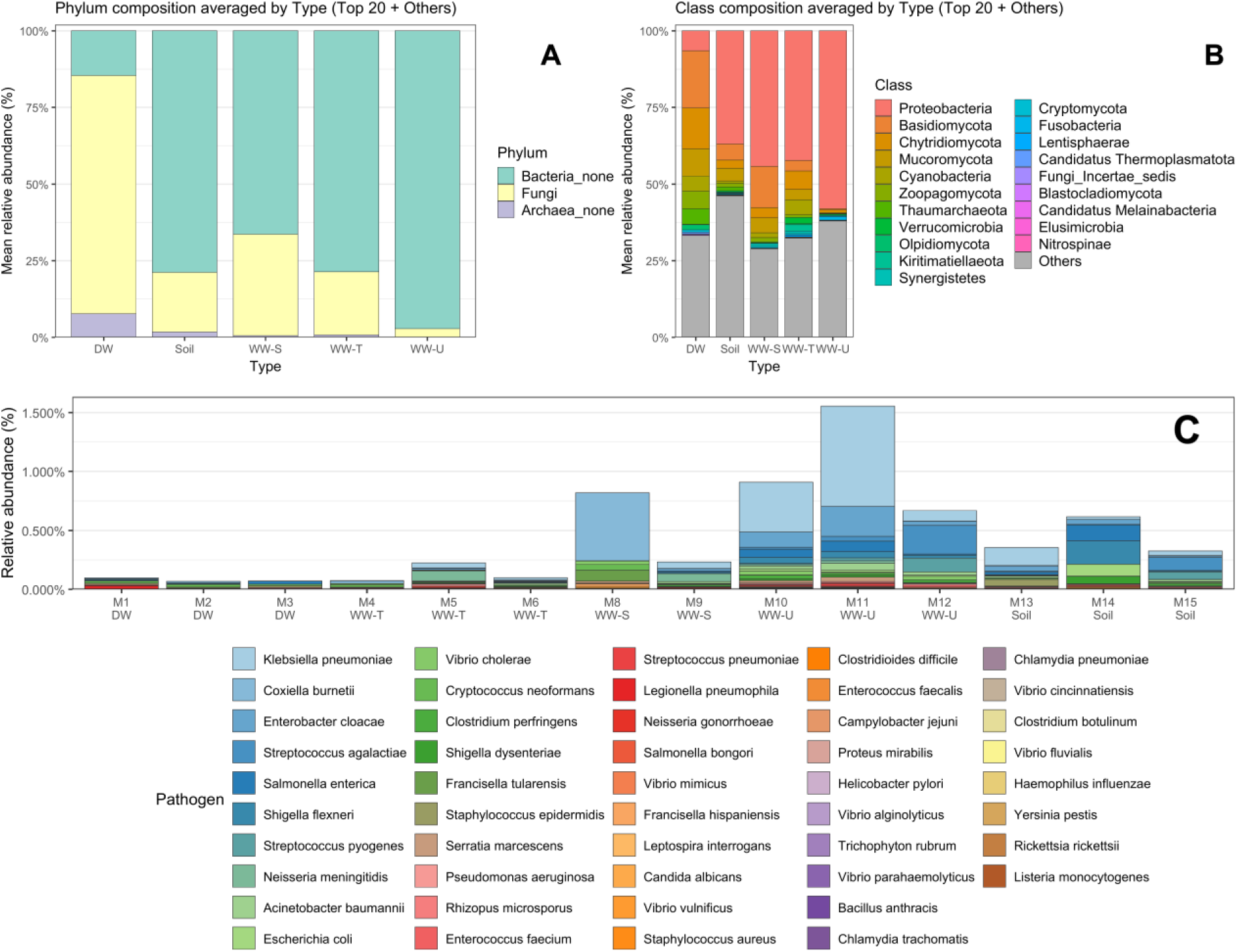
**A.** Mean relative abundance of Bacteria, Fungi and Archaea across environmental sample types. **B.** Mean relative abundance of top 20 microbial classes. **C.** Relative abundance of potential pathogens identified using multiplex metabarcoding (Table S6). Sample types: WW-U – untreated wastewater; WW-S – semi-treated wastewater; WW-T – treated wastewater; DW – drinking water (Table S1).

Class-level profiles (Figure 3B) further differentiated drinking water, soil, wastewater influent/treated (WW-T), and upstream wastewater (WW-U) samples. Notably, drinking water samples contained ∼40% unclassified SSU rRNA reads, reflecting limitations in reference databases and likely representing understudied “microbial dark matter” taxa^42^.

Pathogen distributions varied across sample types. *Klebsiella pneumoniae* was abundant at all sites, particularly in raw or partially treated wastewater, along with *Enterobacter cloacae*, *Shigella dysenteriae*, and *Salmonella enterica* (Figure 3C). Vingungunti raw wastewater exhibited the highest pathogen load, while Mabibo and Msasani sites exhibited slightly lower but comparable abundances. Treated wastewater and drinking water contained markedly fewer pathogen reads, reflecting treatment effects. In contrast, soil from Mabibo unexpectedly contained several pathogenic taxa, including *Shigella flexneri*, *Salmonella enterica*, and *Escherichia coli*, likely due to contamination from nearby wastewater infrastructure.

##### Comparison with metagenomics

Multiplex metabarcoding (PCR-based) was compared with non-enriched ONT metagenomics using sample M11 (WW-U Vingunguti), which had high DNA concentration. The metagenomic run generated ∼1.08 Gb of data (400,800 reads; N50 = 4.22 kb), of which 321,800 reads (467.4 Mb) passed a Q ≥ 8 quality threshold.

Across taxonomic ranks, both approaches showed strong linear agreement in relative taxon abundances (Figure 4A), indicating that multiplex metabarcoding reliably captures dominant community members. However, metabarcoding recovered a broader taxonomic spectrum, identifying 198 unique families in addition to 220 families shared between methods, whereas metagenomics detected 93 unique families not observed in the metabarcoding dataset (Figure 4B).

**Figure 4.**
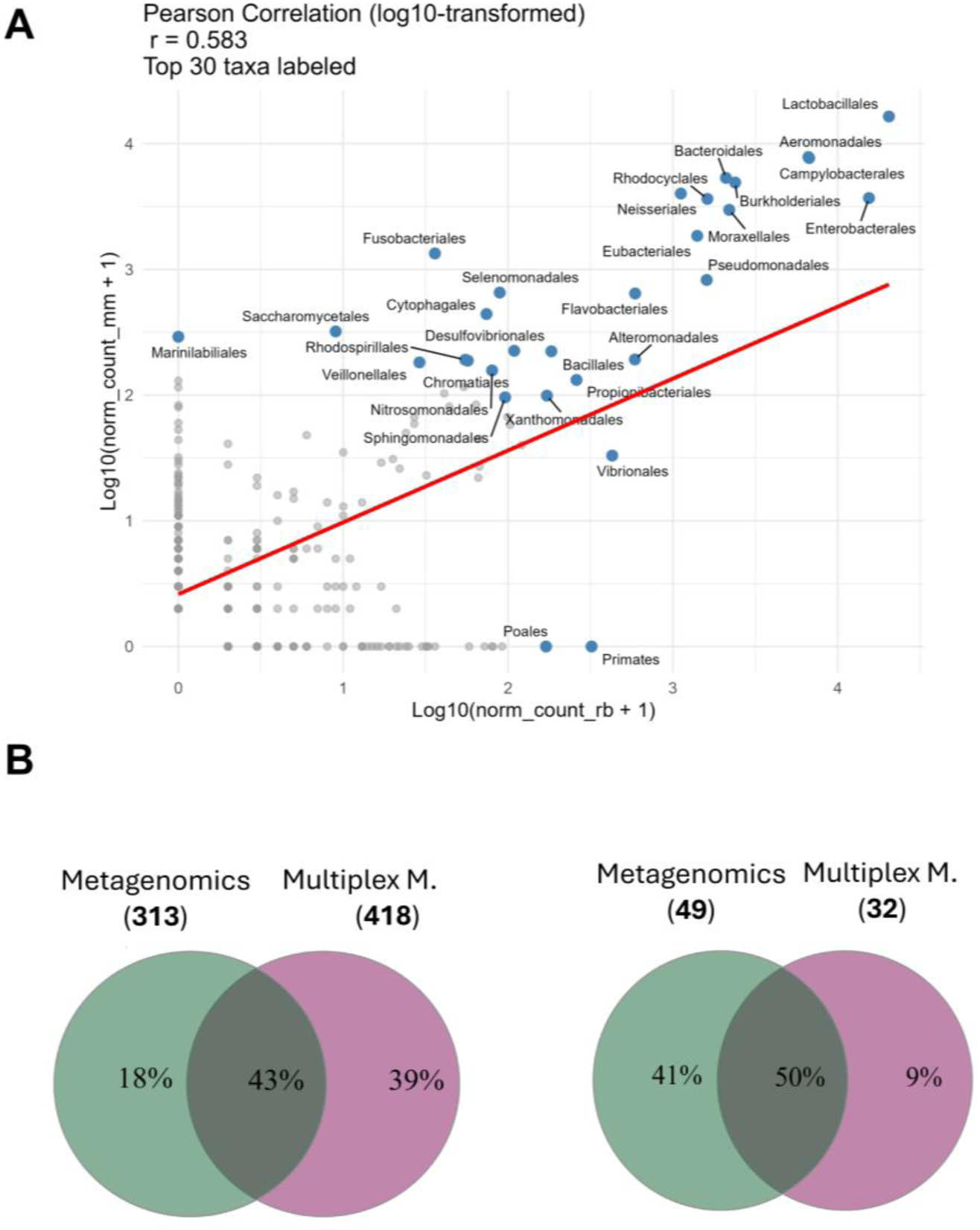
Comparison of bacterial communities between shotgun metagenomics and multiplex metabarcoding for the WW-U Vingunguti (M11) sample: (A) correlation of order-level counts: rb (x-axis) - metagenomic mm (y-axis) multiplex metabarcoding. (B) Venn diagram showing the percentage of shared and unique families (on the left), and Venn diagram the shared targeted pathogenic species (Table S6) (on the right).

#### 3.2.3 EquiPhi-based multiple displacement amplification and shotgun metagenomics

To complement the evaluation presented in Section 3.1.1, which assessed NA extraction for mock community detection, we further tested the Rapid Barcoding Kit combined with EquiPhi-based MDA on the ZymoBIOMICS mock community. Environmental samples from Burkina Faso (Table S1) were also analyzed to evaluate microbial diversity and the distribution of viral sequences and resistomes across four matrices: drinking water, wastewater, surface water, and soil.

##### Microbial diversity

Soil exhibited the highest microbial diversity (Chao1 = 1,473; Shannon = 4.6), followed by wastewater (Chao1 = 1,380; Shannon = 4.2). Drinking water had lower diversity (Chao1 = 1,088; Shannon = 2.8), and surface water the lowest (Chao1 = 646; Shannon = 1.8). In addition, wastewater contained the highest relative abundance for potential human pathogens, including opportunistic species, followed by soil (Figure 5B)

**Figure 5.**
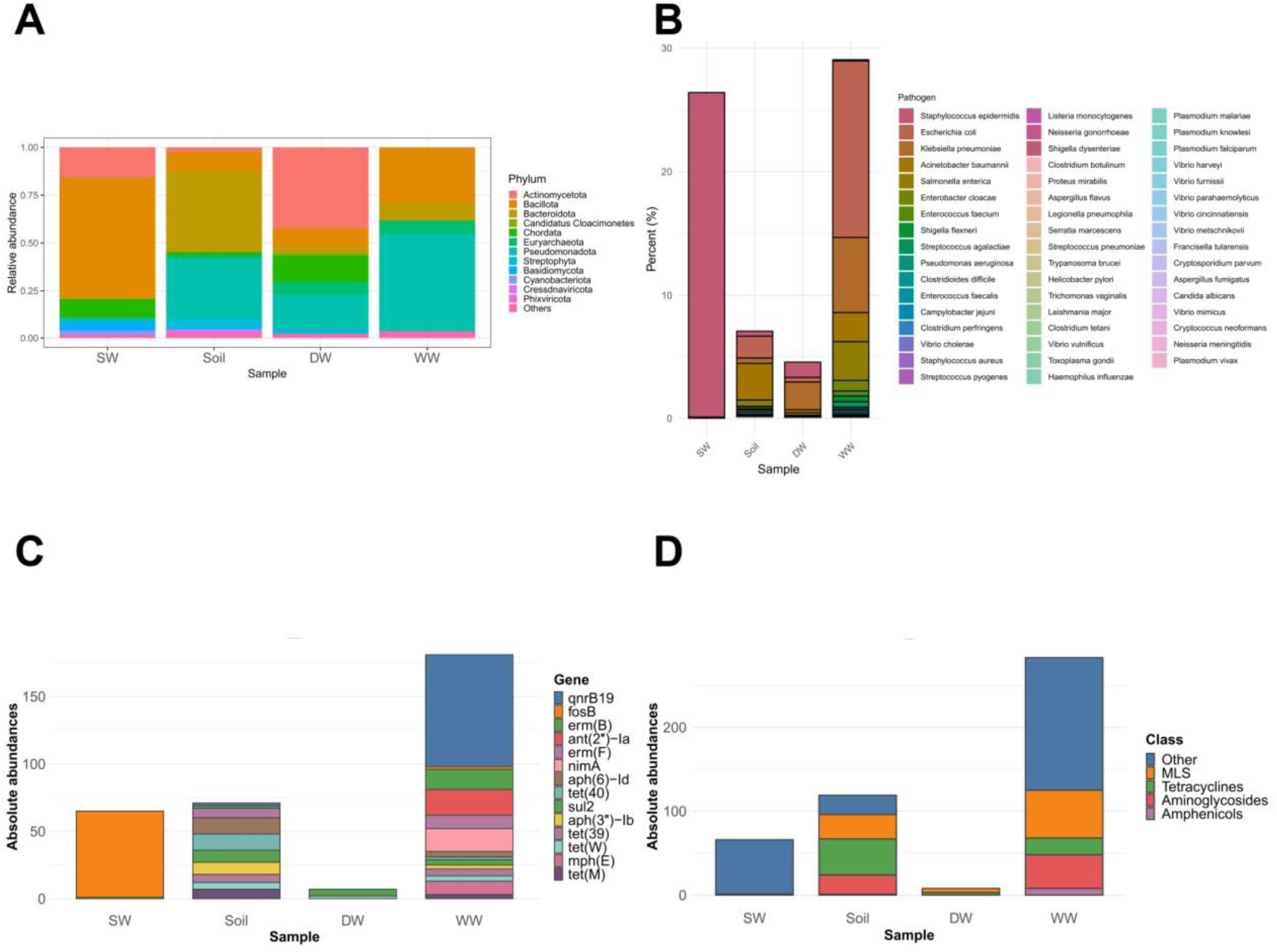
Metagenomic MDA analysis of Burkina Faso environmental samples (Table S1), including surface water (SW), soil, drinking water (DW), and wastewater (WW). A. Relative abundance (%) of microbial taxa at the phylum level. B. Detection of potential human pathogens, including opportunistic species (Table S6). C. Absolute abundance of antibiotic resistance genes (ARGs), showing counts for the top 15 individual ARGs. D. Total ARG counts grouped by resistance class, including MLS (macrolide–lincosamide–streptogramin) resistance.

Surface water was dominated by *Bacillota* (63.5%) and *Actinomycetota* (15.7%), with smaller contributions from *Pseudomonadota* and *Cyanobacteriota*. Eukaryotic sequences included fungi (*Ascomycota* and *Basidiomycota*, ∼4.8%), plant DNA, protists, and human contamination (Figure 5A). Soil was dominated by *Bacteroidota* (43.6%) and *Pseudomonadota* (31.6%), with *Streptophyta* (5.8%) and minor *Bacillota*, *Actinomycetota*, and other bacteria. Archaea, mainly *Euryarchaeota* (1.65%), and low levels of fungi and protists were detected. Drinking water showed high *Actinomycetota* (42.7%) and *Pseudomonadota* (18.1%), with more abundant *Euryarchaeota* (6.7%) than other environments, and minor fungi and plant sequences (*Streptophyta*, 2.3%). Wastewater was dominated by *Pseudomonadota* (51.3%) and *Bacillota* (29.4%), with *Bacteroidota* (8.9%) and other anaerobic/facultative phyla. *Archaea* (*Euryarchaeota*, 7.35%) were notable, while eukaryotic contributions, including human DNA, fungi, and plants, were low. Viral sequences belonged to five classes: *Cressdnaviricota*, *Hofneiviricota*, *Phixviricota*, *Uroviricota*, and Viruses_Incertae_sedis. Soil viruses accounted for 2.44% of classified reads, dominated by *Cressdnaviricota* (1.25%) and *Phixviricota* (1.12%). Minor *Uroviricota* (0.011%) and *Hofneiviricota* (0.061%) were detected, along with trace unclassified viruses. Drinking water viral reads represented 0.34%, largely *Uroviricota* (0.33%), with trace *Cressdnaviricota* (0.0037%) and *Phixviricota* (0.00074%). *Hofneiviricota* and unclassified viruses were nearly absent. Wastewater viruses comprised 0.22% of reads, with *Hofneiviricota* (0.10%) and *Uroviricota* (0.049%) dominant; *Phixviricota* (0.042%) and *Cressdnaviricota* (0.0089%) were also present. Surface water viruses accounted for 0.78% of reads, dominated by *Cressdnaviricota* (0.71%). *Uroviricota* (0.058%) and *Phixviricota* (0.0045%) were minor, with *Hofneiviricota* and unclassified viruses only in trace amounts.

##### AMR

A total of 78 different antibiotic resistance genes (ARGs) were detected in this dataset, spanning the following categories: MLS (macrolides-lincosamides-streptogramins), tetracyclines, aminoglycosides, and amphenicols (Figure 5D). The most abundant gene was qnrB19, classified under “other,” which confers resistance to quinolones. qnrB19, is a plasmid-mediated quinolone resistance gene, was found to be dominant resistome in Burkina Faso samples (Figure 5C). This finding highlights the fluoroquinolones (WHO Essential Medicines) resistance in environmental reservoirs of mobile ARGs in surface water and soil. This may create pathways for resistance to re-enter human populations through different pathways and compromised treatment efficacy. Surface water was dominated by a single ARG, fosB, which encodes a thiol transferase that inactivates fosfomycin and is commonly associated with Gram-positive bacteria. In contrast, the soil sample displayed higher ARG diversity, with no single gene or class dominating. Within the amphenicol category, resistance was detected at only a single read count.

#### 3.2.4 REPLI-g Cell WGA & WTA workflow

The utility of multiplex metabarcoding and REPLI-g approaches for pathogen and ARGs detection was evaluated, including compatibility with standard analysis pipelines such as Epi2meLabs, using environmental samples from Tanzania (Table S1). Overall, REPLI-g consistently detected a higher proportion of pathogens than multiplex metabarcoding, irrespective of DNA or RNA/cDNA starting material (Figure 6C, Figure 7). Mean pathogen detection rates were 6.2% for REPLI-g cDNA, 4.9% for REPLI-g DNA, and 1.1% for multiplex metabarcoding; median values followed the same trend: 6.4%, 5.0%, and 0.9%, respectively. Alpha diversity (Shannon index, Figure 6A) and richness (Chao1 index, Figure 6B) were significantly lower in REPLI-g samples, particularly cDNA (Shannon: p = 0.00116, ANOVA F = 25.587; Chao1: p = 0.00189, ANOVA F = 21.274). Drinking water samples from Mabibo, Vingunguti, and Msasani showed the highest Chao1 and Shannon indices in multiplex metabarcoding, though differences were not statistically significant (Chao1: p = 0.157, ANOVA F = 2.564; Shannon: p = 0.244, ANOVA F = 1.801). Between REPLI-g approaches, DNA samples had higher microbial diversity than cDNA. The relative proportions of *Archaea* and *Eukaryotes* were similar between REPLI-g DNA and multiplex metabarcoding (*Archaea*: 0.06 ± 0.04 vs. 0.07 ± 0.05; *Eukaryotes*: 1.69 ± 0.32 vs. 2.01 ± 1.37). *Bacteria* differed substantially: REPLI-g DNA (54.83 ± 4.71) vs. multiplex metabarcoding (89.40 ± 2.25), with REPLI-g DNA also producing high Viridiplantae sequences (25.29 ± 2.75) absent in metabarcoding.

**Figure 6.**
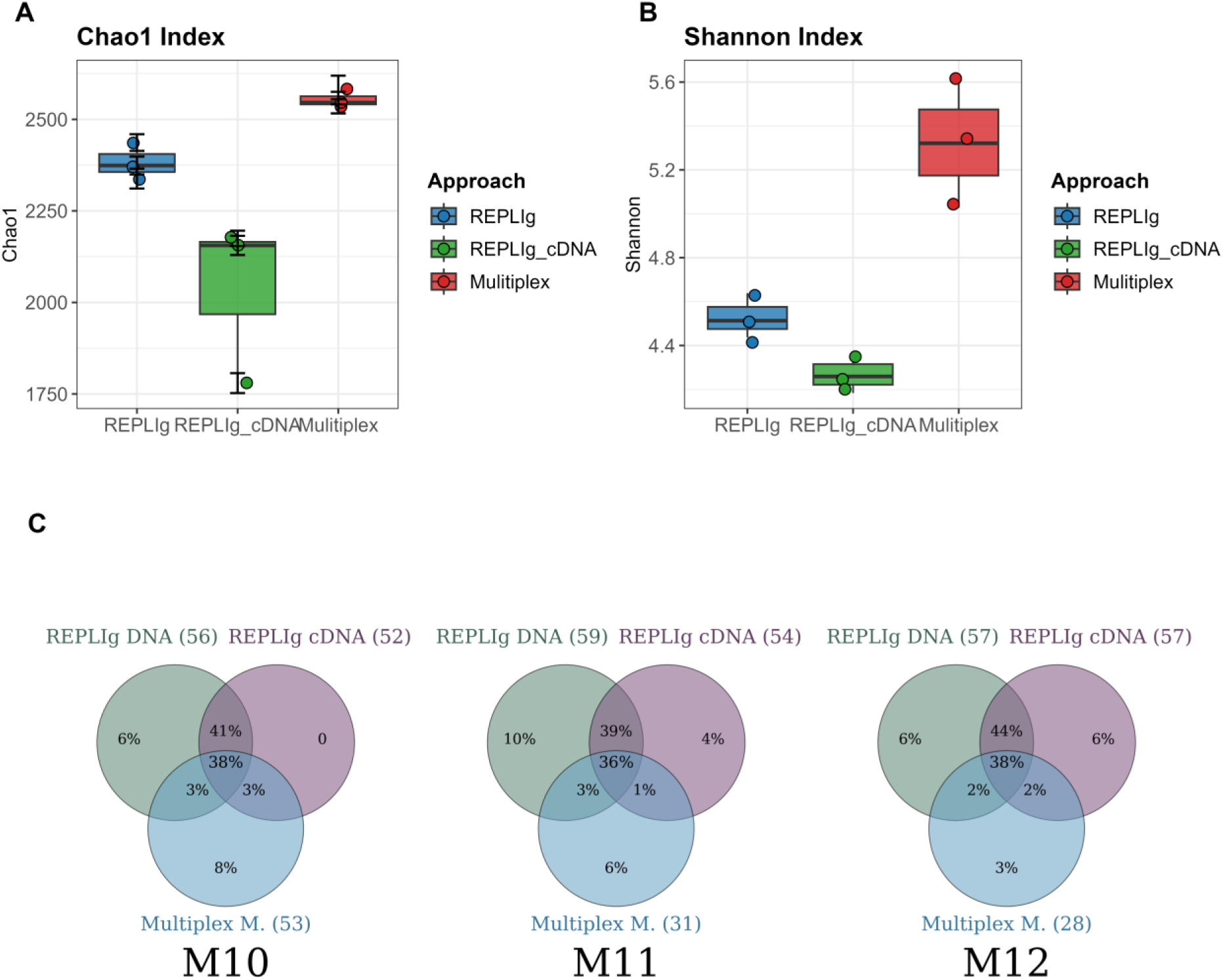
**A** Chao1 index **B** Shanonn index estimates for samples processed with Multiplex Metabarcoding and REPLI-g (DNA and cDNA) protocols. **C** Venn diagrams illustrating the overlap of pathogenic species in three wastewater samples (WW-U: M10, M11, M12) from Mabibo, Vingunguti, and Msasani

**Figure 7.**
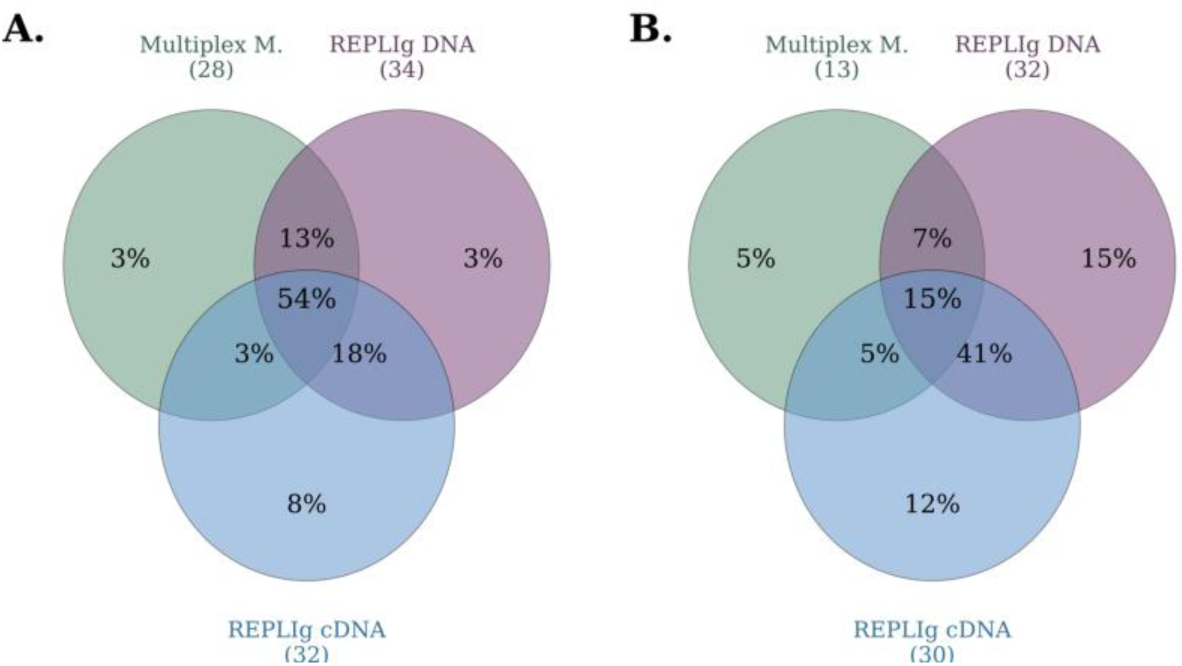
Venn diagrams illustrating the overlap of pathogenic species (Table S6) (A) and their corresponding genera (B) detected in drinking water sample M1 from the Msasani site using REPLI-g DNA, REPLI-g cDNA, and Multiplex Metabarcoding.

##### Viruses

Viruses were a minor portion of reads (REPLI-g DNA: mean 0.0985%, REPLI-g cDNA: 0.00737%, Table 2). REPLI-g DNA mainly contained phages, with fold-changes of ∼13× for viruses and ∼37× for phages compared to cDNA. Non-phage viruses were low in both, including plant- and insect-associated *Virgaviridae* and *Dicistroviridae*; human-associated viruses (*Caliciviridae, Adenoviridae*, *Poxviridae*, *Herpesviridae*) were rare.

**Table 2.** Contribution (%) of viral sequencing reads across environmental samples in the dataset.

| Category/Sample | REPLIg_DNA |  |  | REPLIg_cDNA |  |  |
| --- | --- | --- | --- | --- | --- | --- |
|  | 10M | 11M | 12M | 10M | 11M | 12M |
| Viruses (%) | 0.0860 | 0.1225 | 0.0871 | 0.0063 | 0.0099 | 0.0059 |
| Non-phage viruses (%) | 0.0047 | 0.0102 | 0.0062 | 0.0039 | 0.0064 | 0.0044 |
| Phage viruses (%) | 0.0813 | 0.1123 | 0.0810 | 0.0024 | 0.0035 | 0.0015 |

##### Antimicrobial Resistance (AMR)

The REPLI-g workflow did not detect ARGs in WW-U and DW samples using Epi2me Labs, though ARGs were confirmed in Vingunguti WW-U via shotgun metagenomics of non-enriched DNA. The resistome was dominated by genes conferring resistance to aminoglycosides, macrolides, sulfonamides, trimethoprim, β-lactams, fluoroquinolones, and tetracyclines. The five most abundant ARGs were *sul*1 (23 reads), *aph*(6)-Id (14), *msr*(D) (13), *dfr*A1 (10), and *aph*(3’)-Ib (9), alongside sul2, mph(E), msr(E), and multiple β-lactamases (blaOXA, blaCARB, blaGES, blaMOX, cfxA). Genes conferring resistance to chloramphenicol, rifampicin, colistin (mcr variants), and multidrug efflux pumps (oqxAB) were also detected, revealing a diverse resistome spanning multiple mechanisms.

##### Availability

Sequencing datasets are publicly available in the NCBI SRA under BioProject PRJNA1259765 (amplicon datasets SRR33650859–SRR33651847; other datasets SRR33652213–SRR33652358 and SRR33454201–SRR33471298).

### 3.3 Selection, and integration of qPCR assays

#### 3.3.1 Implementation and optimization of qPCR reactions

qPCR assays exhibited amplification efficiencies ranging from 0.78 to 1.22, with 10 of the 12 assays demonstrating efficiencies between 0.84 and 1.12 (Figure S5 and Figure S6). The assays targeting *Vibrio cholerae* (*ctxA*) and wild poliovirus type 1 (WPV1) exhibited the most extreme efficiency estimates (Figure S5). The published annealing temperatures for *ctx*A and WPV1 assays were 63 °C and 50 °C, respectively, differing from the common 60 °C annealing temperature used here. These differences are likely to reflect suboptimal thermocycling conditions rather than limitations of the Biomeme qPCR platform.

#### 3.3.2 Pilot testing of field-collected samples

Among the eight assays applied to field-collected environmental samples collected in Tanzania, three targets: *Klebsiella pneumoniae*, *Vibrio cholerae*, and Rotavirus were detected predominantly in wastewater samples (Table S8).

## 4 Discussion

This study validates a ML molecular workflow for WES within the ODIN consortium, deployed in Tanzania, and Burkina Faso. Workflow performance was evaluated through benchmarking of nucleic acid extraction against the widely used ZymoBIOMICS DNA Miniprep Kit and by head-to-head comparison of sequencing strategies, including multiplex metabarcoding, MDA-based shotgun metagenomics, and REPLI-g WGA/WTA. The workflow was explicitly designed for field deployment, minimizing reliance on high-power centrifugation, permanent bead-beating equipment, and cold-chain logistics, while incorporating structured training to reduce operator-dependent variability (https://odin-wsp.tghn.org/). A substantial portion of method validation was performed under controlled laboratory conditions, enabling rigorous assessment of analytical performance. While this approach strengthens methodological robustness, it does not fully capture operational constraints in field settings. Nevertheless, successful application to pilot environmental samples from Tanzania and Burkina Faso supports its transferability to real-world contexts.

The nucleic acid extraction workflow, based on chemical lysis combined with magnetic bead-based purification, performed consistently across wastewater, sediment, mock communities, and spiked field samples, yielding DNA of sufficient quantity, purity, and representativeness. Compared with the ZymoBIOMICS kit, performance was comparable or slightly improved in terms of DNA integrity, while avoiding dependence on cold-chain storage and fixed laboratory infrastructure. These features make the workflow particularly suitable for low-resource settings in sub-Saharan Africa, where logistical constraints and infrastructure variability remain major barriers to routine molecular surveillance^1,43^. Chemical lysis combined with magnetic bead–based purification is a well-established method used in many studies^44–46^, and we again demonstrate its suitability for field deployment in a mobile laboratory.

ONT sequencing enabled flexible application of both metabarcoding and shotgun metagenomics^15,18,47^. Metabarcoding provided a rapid and cost-effective approach for routine monitoring of dominant taxa^19,20,48^, while shotgun sequencing offered higher taxonomic and functional resolution, including detection of low-abundance taxa, viruses, and antimicrobial resistance genes^24,26,49^. These complementary strengths support a tiered surveillance strategy in which metabarcoding is used for high-frequency screening and shotgun metagenomics is applied selectively for in-depth characterization.

A key advancement of this study is the implementation of a cost- and time-efficient multiplex metabarcoding strategy enabling simultaneous amplification of SSU rRNA genes across bacteria, archaea, and eukaryotes in a single run. This approach reduces laboratory complexity and cost compared with domain-specific workflows and minimizes ONT-related length bias by standardizing amplicon sizes. Inclusion of archaea, while not directly linked to pathogenicity, enhances ecological resolution and broadens interpretation of microbial dynamics in engineered and natural water systems.

Method concordance analyses in Tanzanian drinking water and untreated wastewater demonstrated moderate to high agreement between metabarcoding and qPCR for key pathogens. For *Vibrio cholerae*, concordance was 66.7%, with metabarcoding detecting *Vibrio* spp. even in qPCR-negative samples, highlighting its broader detection scope. However, qPCR provided essential specificity for virulence-associated markers such as *ctxA*, confirming pathogenic potential that sequencing alone could not resolve. For *Klebsiella pneumoniae*, higher agreement (83.3%) was observed, consistent with improved detection of abundant taxa. In contrast, REPLI-g WGA/WTA showed lower concordance and greater variability, with additional detections likely reflecting amplification bias or stochastic effects in low-template samples.

Overall, metabarcoding showed closer agreement with qPCR than REPLI-g-based approaches, although both sequencing methods detected additional low-abundance taxa not captured by qPCR. These findings reinforce the complementary roles of sequencing and targeted molecular assays: sequencing provides broad, hypothesis-free organism detection, while qPCR delivers high sensitivity and functional specificity for clinically relevant markers^50^. A two-tier strategy—untargeted sequencing followed by targeted qPCR confirmation—therefore improves diagnostic confidence, reduces false positives and negatives, and aligns with current recommendations for environmental pathogen surveillance^51,52^. Equipping mobile laboratories with both sequencing platforms and flexible qPCR assay panels is therefore critical for rapid, reliable pathogen detection and characterization in field settings^53^.

Despite these advances, viral pathogen detection remains a major limitation in metagenomic workflows^54^. The absence of universal viral markers, coupled with high sequence diversity, limits comprehensive detection using standard approaches. Although multiple displacement amplification (MDA) can enrich viral nucleic acids, it introduces technical challenges including pore clogging and formation of branched DNA structures that may reduce ONT sequencing efficiency. Enzymatic treatments such as endonuclease T7 partially mitigate these effects but do not fully resolve them^55^. Consequently, lower sequencing yields were observed in some runs, although they remained sufficient to recover all species in mock communities.

Among amplification strategies, REPLI-g WGA/WTA proved less suitable for field deployment due to high cost, operational complexity, and artefact formation, particularly chimeric sequences arising during ligation steps that complicate downstream bioinformatic interpretation. In contrast, Phi29-based WGA with 5′-phosphorylated random hexamers, combined with optimized enzymatic treatments, provided a more robust and field-compatible solution for DNA-based pathogen and resistome surveillance. This approach performed reliably across diverse environmental matrices, including low-biomass drinking water samples, without requiring large-volume filtration. Our previous work showed that MDA using the EquiPhi29 kit with 5′-phosphorylated random hexamers effectively captures extensive ARG diversity and overall microbial composition in wastewater, even when using the Rapid Barcoding kit^26^.

However, RNA virus detection remains unresolved within this workflow and requires dedicated, highly sensitive methods such as targeted RT-qPCR. This highlights a persistent gap in comprehensive environmental virology, even within advanced metagenomic frameworks.

Overall, these results emphasize that no single method is sufficient for comprehensive environmental surveillance. Instead, optimal performance is achieved through method integration tailored to analytical goals. The optimized ML workflow provides a practical balance between cost, portability, and analytical resolution, making it suitable for deployment in resource-limited settings. Future work should focus on improving viral detection, reducing amplification bias, and integrating sequencing and qPCR platforms into unified One Health surveillance systems capable of supporting rapid public health decision-making.

## 5 Conclusion

This study validates a field-deployable ML workflow for WES of pathogens and AMR. The results show that optimized, low-equipment protocols can generate reliable, decision-grade data in SSA when extraction and library preparation are adapted to sample type, amplification biases in shotgun metagenomics are addressed, and viral detection is supported by sensitive targeted assays. The integrated use of metabarcoding, shotgun metagenomics, and qPCR provides a cost-effective and scalable One Health approach for environmental monitoring.

Future work should focus on standardized viral detection workflows, automation at the point of sampling, and strengthened data infrastructure, workforce capacity, and governance to enable timely public health action. Further evaluation of cost efficiency and large-scale field performance is also required to fully establish operational feasibility and public health impact.

## Data Availability

All data produced in the present study are available upon reasonable request to the authors

## Acknowledgement

This study was supported by the European Commission H2020 and EDCTP3 through the ODIN project (grant no. 101103253, *Strengthening Environmental Surveillance to Advance Public Health Action*) and the ODIN-Mpox project (grant no. 101195186, *Implementing wastewater and environmental surveillance for Mpox in sub-Saharan Africa*). Additional support was provided by the SusOffAqua project (Research Council of Norway, project no. 328724) for all work involving sediment samples. Mpox virus material was supplied by the European Virus Archive Global (EVA-GLOBAL), funded under the European Union’s Horizon 2020 research and innovation programme (grant agreement no. 871029).

We would also like to thank tool developers (Samuel Wilkinson, Áine O’Toole) for their support in troubleshooting the Mpox bioinformatic pipeline.

## Author contributions

**Andrea Bagi:** Investigation, data curation, formal analysis, writing – original draft, review & editing. **Ananda Tiwari:** Writing – original draft, review & editing. **Chikwendu Chukwudiegwu Mbachu:** Formal analysis, investigation, writing – original draft. **Dylan Shea:** Formal analysis, investigation, data analysis, data curation, writing – original draft, review & editing. **Tam T. Tran:** Formal analysis, investigation, data analysis, data curation, writing – original draft, review & editing. **Christian Tahita:** Funding acquisition, writing-editing **Palpouguini Lompo:** Sampling, investigation, writing- editing **Peter Mkama:** Sampling, investigation. **Eric Lyimo:** Sampling, investigation, writing- editing **Vito Baraka:** Funding acquisition, writing – review & editing. **Alan Le Tressoler:** Sampling, Investigation. **Tarja Pitkänen:** Funding acquisition, writing – review & editing. **Adriana Krolicka:** Conceptualization, funding acquisition, project administration, supervision, investigation, data curation, formal analysis, visualization, writing – original draft, review & editing.

