## Supplementary Materials for "Mobile wastewater and environmental surveillance in low-resource settings: validating an integrated laboratory workflow for pathogen detection"

### Supplementary Figures

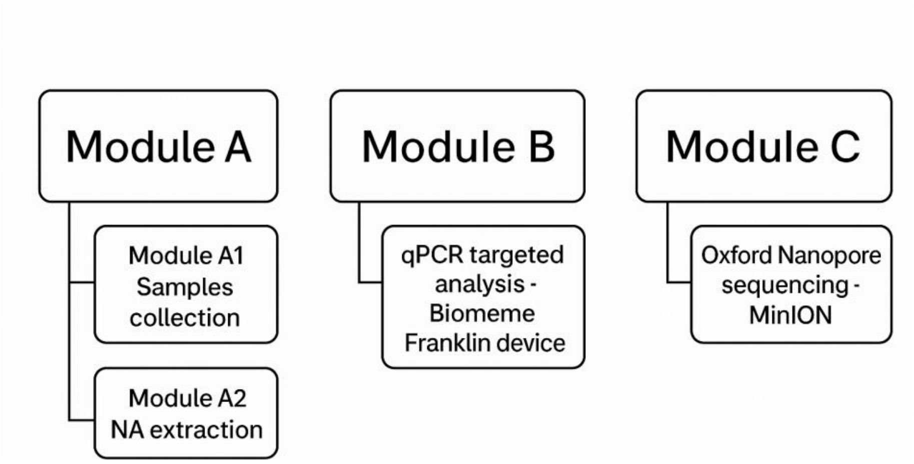


**Figure S1.** The envisioned mobile laboratory with a modular design.

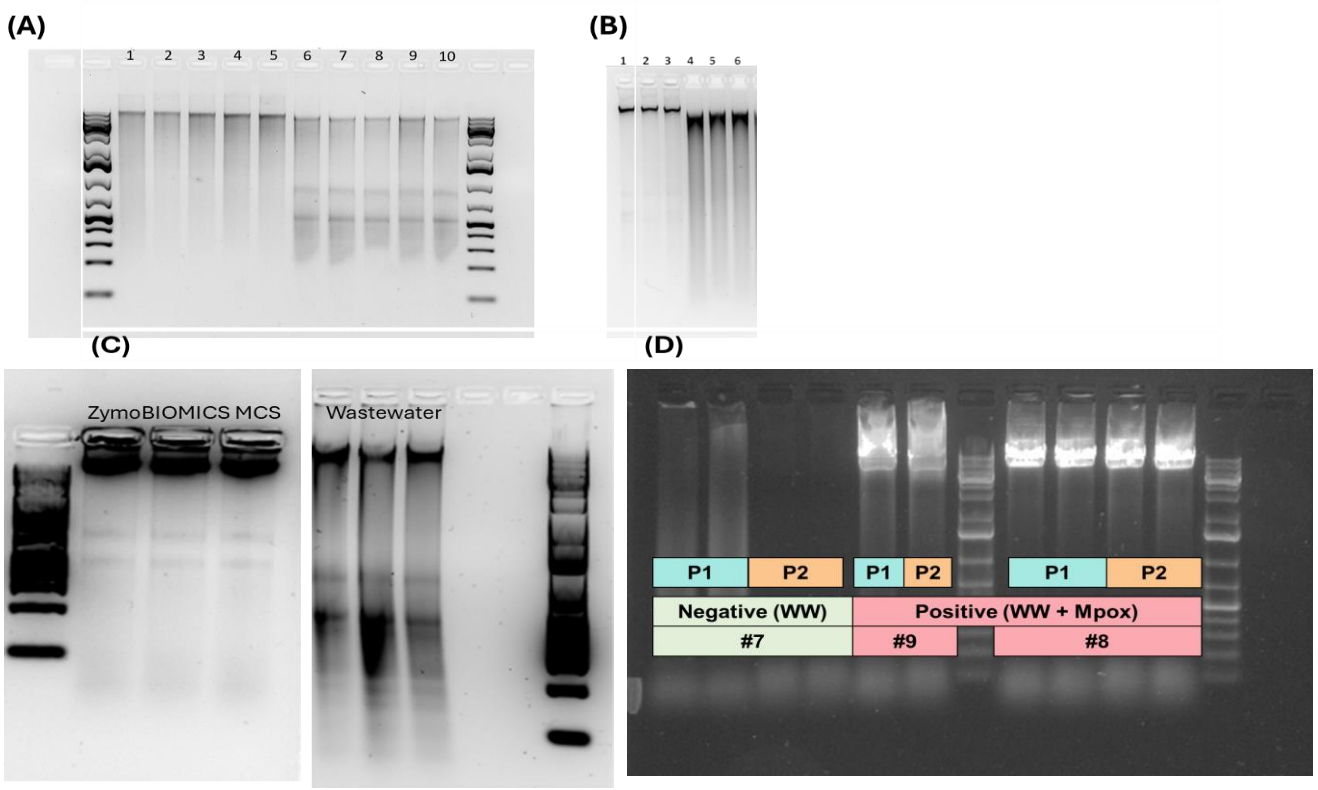


**Figure S2.** Agarose gel electrophoresis of total nucleic acids and PCR amplification results. (A) Total nucleic acids extracted from wastewater (WW). Lanes 1–5: ZymoBIOMICS DNA Miniprep Kit; lanes 6–10: ML nucleic acid extraction method. (B) Total nucleic acids extracted from sediment samples. Lanes 1–3: ML extraction method; lanes 4–6: ZymoBIOMICS DNA Miniprep Kit. (C) ZymoBIOMICS mock community spiking experiment. Left: DNA extracted from the ZymoBIOMICS Microbial Community Standard (MSC); right: DNA extracted from WW spiked with MSC. The ML workflow was validated using the mock community. (D) PCR amplification of extracted DNA using primer pools. Gel image showing products amplified with either Pool 1 (P1) or Pool 2 (P2) primers

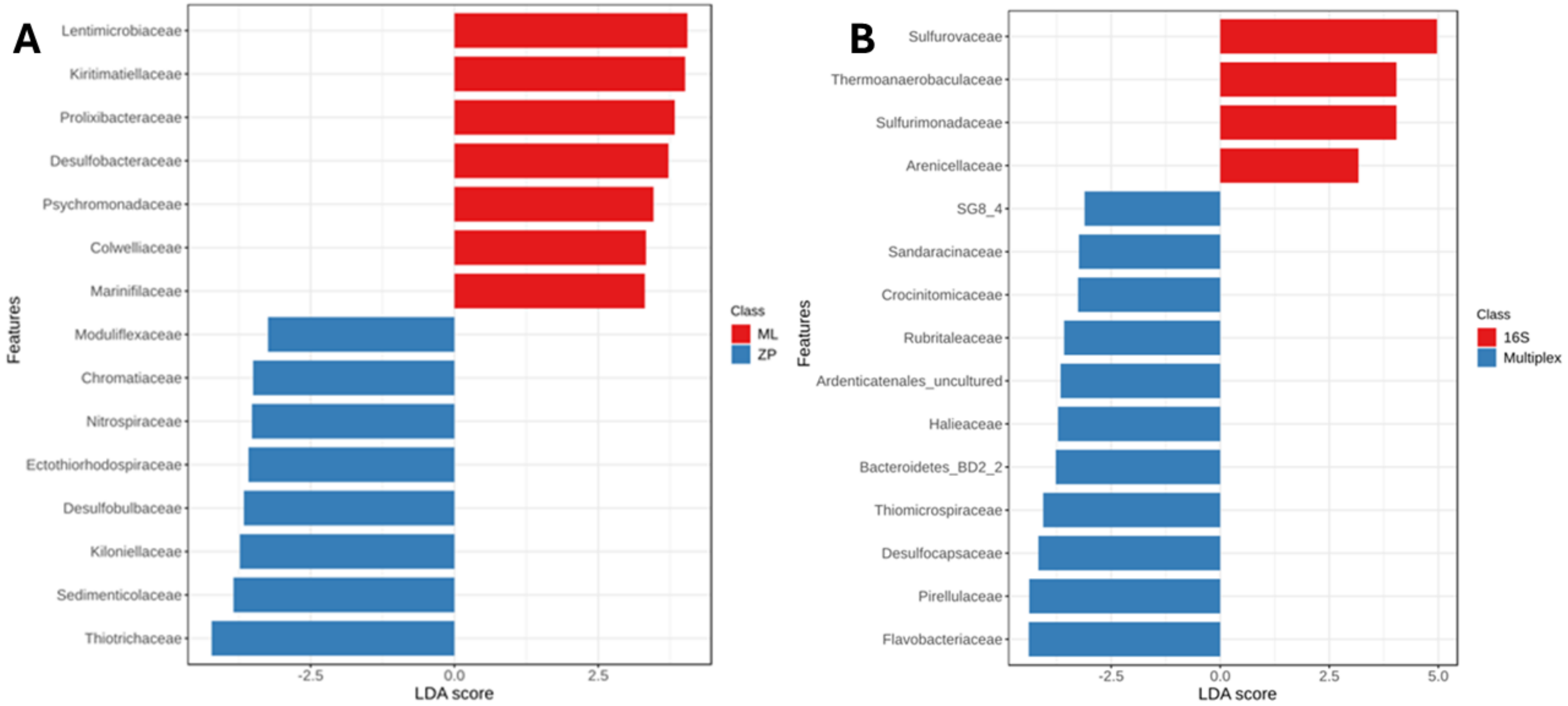


**Figure S3** LEfSe analysis of taxa enriched by library preparation method **A.** Distinct microbial groups were associated with the ML NA extraction and the ZymoBIOMICS kit (FDR-adjusted $p < 0.05$). **B.** Distinct microbial groups were associated with the 16S kit (Oxford Nanopore) and the Multiplex primer set, comparing ZP_Multiplex (Zymo) and ML_Multiplex (Mobile Lab) protocols.

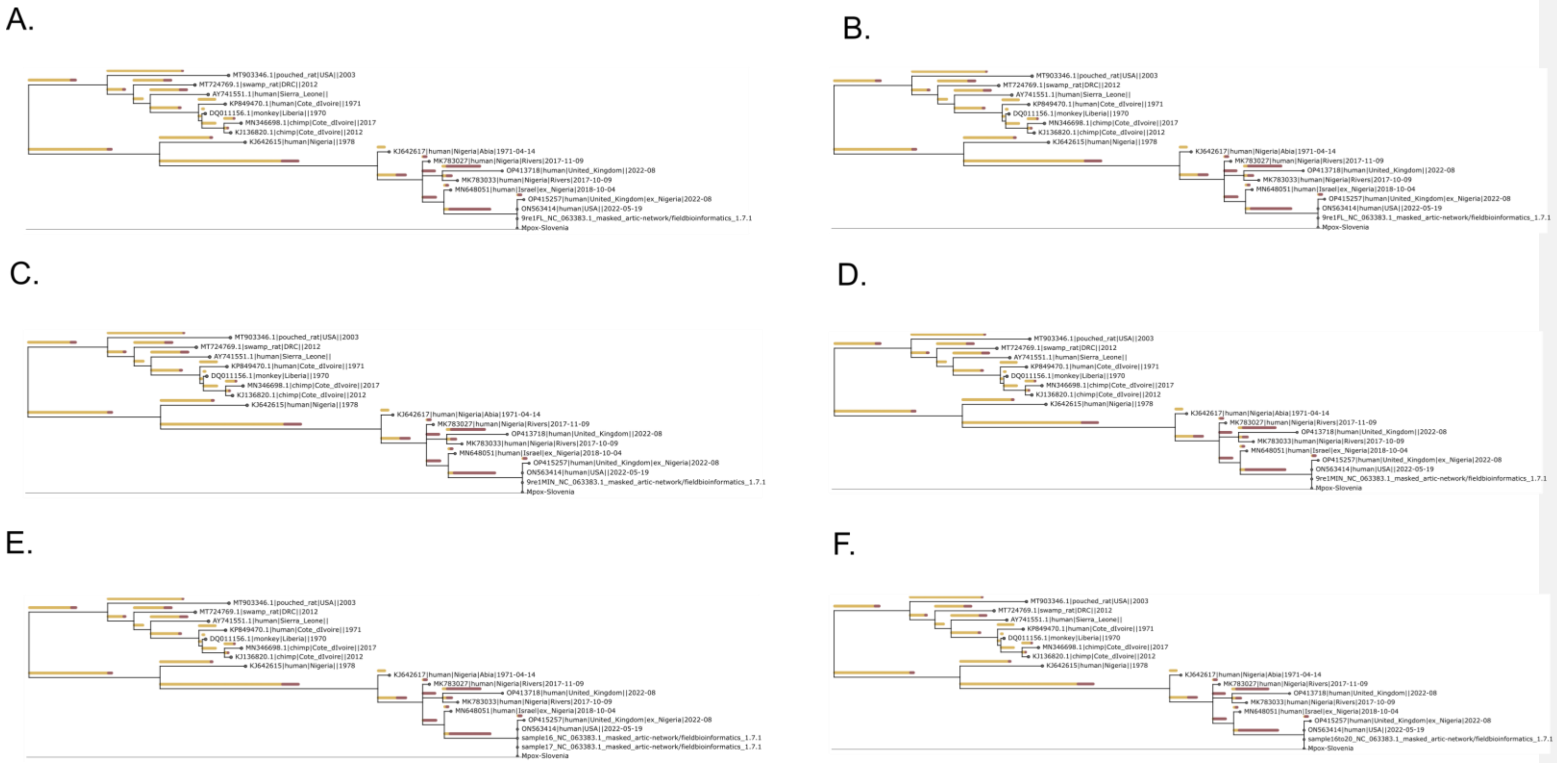


**Figure S4.** Phylogenetic consistency of Mpox virus sequencing across platforms, primer schemes, and data volumes. Identical phylogenetic trees were obtained from a Mpox-spiked wastewater sample sequenced using Flongle (A, B) or MinION (C, D) flowcells, processed with the *artic-mpxv-nf* workflow using either the default primer scheme (A, C) or a custom primer scheme (B, D), and generated from different output data volumes (E: 4.5 MB; F: 200 MB). The Mpox Slovenia reference sequence was provided by the supplier and is publicly available via the European Virus Archive (https://www.european-virus-archive.com/virus/monkeypox-virus-2022-slovenia-ex-gran-canaria-inactivated-virus).

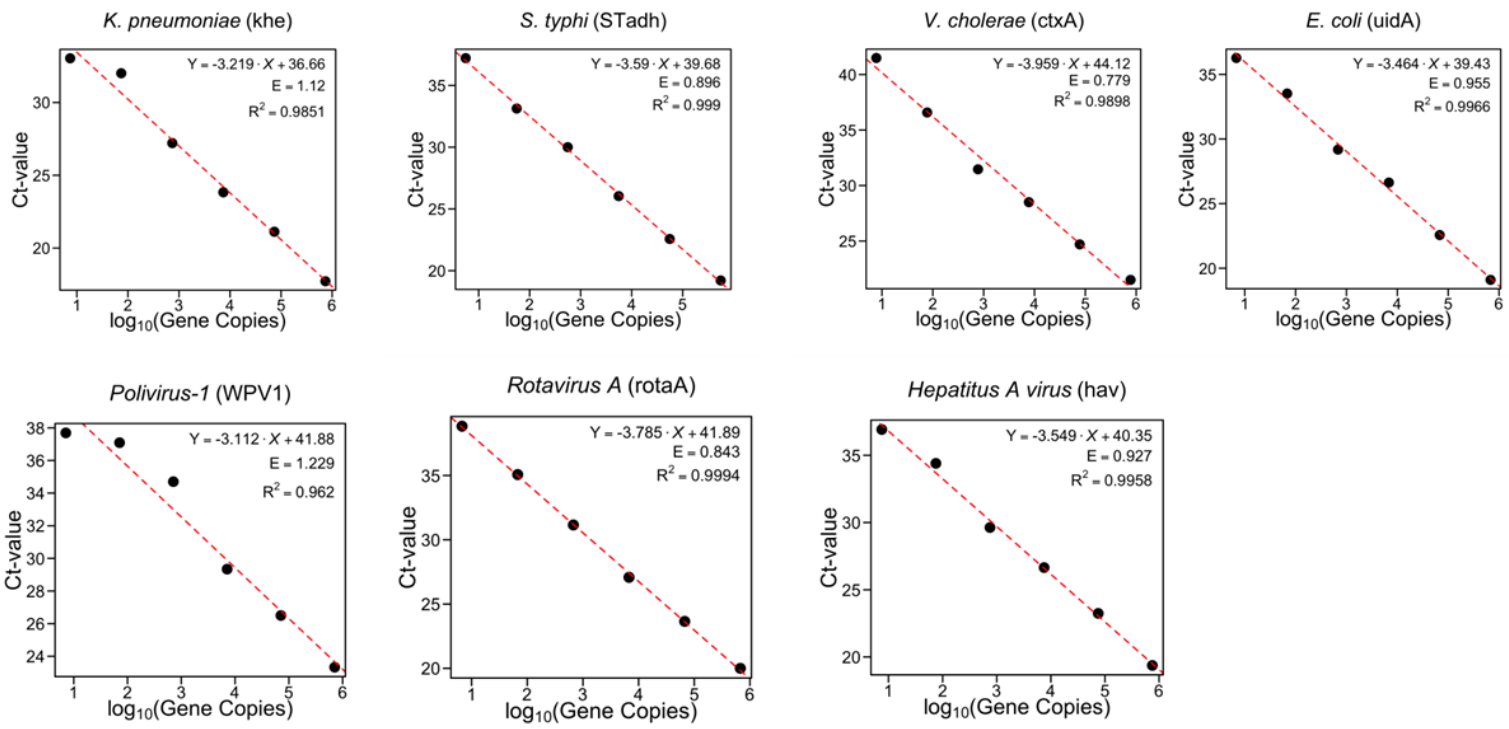


**Figure S5.** Ct-values as a function of gene copy concentrations for 10-fold standard dilutions. The equation for the relationship between Ct-values and gene copies (log10) are printed in the plot area and also depicted visually using a red dotted line. Efficiency estimates (E) and R-squared values ($R^2$), calculated from linear regression results, are also printed inside the plot area.

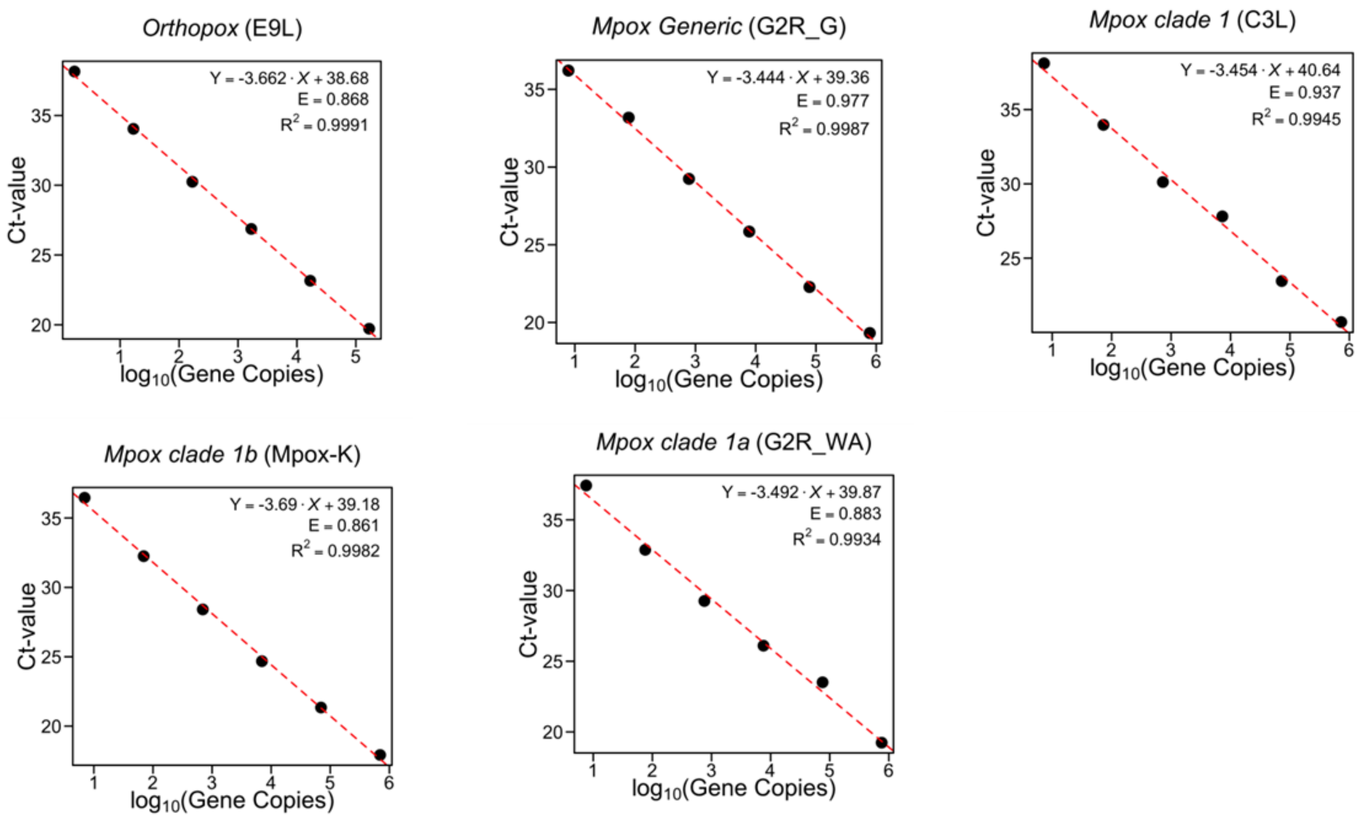


**Figure S6.** Ct-values as a function of gene copy concentrations for 10-fold standard dilutions. The equation for the relationship between Ct-values and gene copies (log10) are printed in the plot area and also depicted visually using a red dotted line. Efficiency estimates (E) and R-squared values ($R^2$), calculated from linear regression results, are also printed inside the plot area.

### Supplementary Tables

**Table S1.** Overview of samples. SE1: wastewater sample used for DNA spiking with ZymoBIOMICS Standard Community, SE2: wastewater sample used for Mpox spiking experiment

| | | | | | NA extraction validation | | | Sequencing approaches | | | Targeted approach |
|---|---|---|---|---|---|---|---|---|---|---|---|
| **Coordinates** | **Country** | **Sample type** | **Sample name** | **Site/ Localization** | **ML vs Zymo** | **Mock community** | **MPXV capture** | **Multiplex metabarcoding** | **MDA/ONS metagenomics** | **REPLI-g WGA/WTA** | **qPCR imp. Biomeme** |
| **59.020073 N,5.620413 E** | NO | WW_U | WW-Mek | Mekjarvik | x | x (SE1) | | | x | | |
| **69.670000 N,18.970000 E** | NO | WW_U | WW-Tro | Tromsø | | | x(SE2) | | | | |
| **59.065186N,5.412999E** | NO | Sed | Sed-Kv | Kvitsøy | x | | | | | | |
| **6.8380 S,39.2358 E** | TZ | DW | M1 | Msasani | | | | x | | x | x |
| **6.7656 S,39.2525 E** | TZ | DW | M2 | Vingunguti | | | | x | | | x |
| **6.8094 S,39.2245 E** | TZ | DW | M3 | Mabibo | | | | x | | | x |
| **6.8094 S,39.2245 E** | TZ | WW_T | M4 | Mabibo | | | | x | | | x |
| **6.7656 S,39.2525 E** | TZ | WW_T | M5 | Vingunguti | | | | x | | | x |
| **6.8380 S,39.2358 E** | TZ | WW_T | M6 | Msasani | | | | x | | | x |
| **6.8380 S,39.2358 E** | TZ | WW_S | M7 | Msasani | | | | x | | | x |
| **6.8094 S,39.2245 E** | TZ | WW_S | M8 | Mabibo | | | | x | | | x |
| **6.7656 S,39.2525 E** | TZ | WW_S | M9 | Vingunguti | | | | x | | | x |
| **6.8380 S,39.2358 E** | TZ | WW_U | M10 | Msasani | | | | x | | x | x |
| **6.7656 S,39.2525 E** | TZ | WW_U | M11 | Vingunguti | | | | x | | x | x |
| **6.8094 S,39.2245 E** | TZ | WW_U | M12 | Mabibo | | | | x | | x | x |
| **6.8380 S,39.2358 E** | TZ | Soil | M13 | Msasani | | | | x | | | x |
| **6.7656 S,39.2525 E** | TZ | Soil | M14 | Vingunguti | | | | x | | | x |
| **6.8094 S,39.2245 E** | TZ | Soil | M15 | Mabibo | | | | x | | | x |
| **12.3828 N, 1.5118 W** | BF | DW | DWBF | Ouagadougou | | | | | x | | |
| **12.3861 N, 1.5027 W** | BF | WW | WWBF | Ouagadougou | | | | | x | | |
| **12.3866 N, 1.5259 W** | BF | SW | SWBF | Ouagadougou | | | | | x | | |
| **12.2229 N, 1.5834 W** | BF | Soil | SoBF | Kilwin/Tanghin Ouagadougou | | | | | x | | |

**Table S2.** Primer sequences used in the multiplex metabarcoding .

| Domain | Oligo name | Primer Sequence (5’ ® 3’) | Size of amplicon (bp) | Reference |
|---|---|---|---|---|
| Bacteria | Bac27F<br>Bac1492R | AGRGTTYGATYMTGGCTCA<br>GRGYTACCTTGTTACGACTT | 1465 | Callahan et al., 2019 |
| Archaea | Arch1F<br>Arch1492R | TCCGGTTGATCCYGCBRG<br>CGGNTACCTTGTKACGAC | 1492 | Bahram et al., 2019 |
| Eukaryotes | Euk83F<br>Euk1623R | GAAACTGCGAATGGCTC<br>GGGCGGTGTGTACAARGRG | 1540 | Amaral-Zettler et al., 2009 |

**Table S3.** Primer sequences used for MPXV amplicon sequencing, including primer names, sequences, target regions, and primer pool assignment. *Mpox virus spiking experiment assessing MPXV capture and ML nucleic acid extraction performance.*

| **Name** | **Pool** | **Sequence (5'-3')** |
|---|---|---|
| MPXV_Seq_1_R | 1 | ACCAAGCTAAGCGACTACCATC |
| MPXV_Seq_5_F | 1 | CGGAAGGGACTATATGACTAACTTATCT |
| MPXV_Seq_7_R | 1 | ACTATGGATCCCCACCACTTGA |
| MPXV_Seq_11_F | 1 | CGAAGATCGTTGCCAGCAAC |
| MPXV_Seq_13_R | 1 | CATTTTCTCAGACTGGATAAATGGC |
| MPXV_Seq_17_F | 1 | GTGCACATAAACCTCTGGCACT |
| MPXV_Seq_19_R | 1 | TCGTTTGTTGGAAGGGCAGAAA |
| MPXV_Seq_23_F | 1 | ACGCGTTCACTATCTCCAGAGA |
| MPXV_Seq_25_R | 1 | AGATCAAAACCAGCTGCGATGT |
| MPXV_Seq_29_F | 1 | GCCCCAGGATCTCCAACAAATT |
| MPXV_Seq_31_R | 1 | GGAAATTAAACGGCGCCAATGT |
| MPXV_Seq_35_F | 1 | TGACCCTCACTCCGATATCTGG |
| MPXV_Seq_37_R | 1 | CGGGTTCAGAAATATCG |
| MPXV_Seq_41_F | 1 | AACCATAGACGCCAACGAATCC |
| MPXV_Seq_43_R | 1 | AGTTTCCAGACTGGTATCCGAGT |
| MPXV_Seq_47_F | 1 | ACGGCGATTTCGTAGAAGTTGA |
| MPXV_Seq_49_R | 1 | GAAGGAGGCCTTCGTTCGAAAT |
| MPXV_Seq_53_F | 1 | TTGACGATGATGATTGATCACTATTACAC |
| MPXV_Seq_55_R | 1 | CCAACACTACATTTGAGGTGCG |
| MPXV_Seq_59_F | 1 | GTAAACGGGTGTCATGTGACGA |
| MPXV_Seq_61_R | 1 | TCACGAAAGAAGGATGTCTACCG |
| MPXV_Seq_65_F | 1 | TCCCTCTTCTTCTACCTTTCCCA |
| MPXV_Seq_67_R | 1 | TCGAGTCATTTTACGCACGGTT |
| MPXV_Seq_71_F | 1 | GAATCTCTAACGCTGGCCTCTG |
| MPXV_Seq_2_F | 2 | ACGGCTTCTGTATTCGTTGTCT |
| MPXV_Seq_4_R | 2 | ACAACTTCATAGCAAATTCCACATTCT |

| | | |
|---|---|---|
| MPXV_Seq_8_F | 2 | AGGCTATGTTTCGCCCATCATC |
| MPXV_Seq_10_R | 2 | GCGGTGTTCCACACTCTGTATT |
| MPXV_Seq_14_F | 2 | CCTCCGCATACGCATTACAGTT |
| MPXV_Seq_16_R | 2 | TGGTACGCGTAATAATATGCCGA |
| MPXV_Seq_20_F | 2 | ACGCGTATGGATGGATACCAGA |
| MPXV_Seq_22_R | 2 | TGTTTCGACTGGAGAATCATCCA |
| MPXV_Seq_26_F | 2 | CCCACATCATTTCAACCAAAGACG |
| MPXV_Seq_28_R | 2 | TGGTGGATGTGAACAGCATACG |
| MPXV_Seq_32_F | 2 | GAGGATATGGTGGTCGACGGAT |
| MPXV_Seq_34_R | 2 | TCGTAATCTATGCCTTTAGCTAGAGC |
| MPXV_Seq_38_F | 2 | CCACATATGATAAACGGCAAACCA |
| MPXV_Seq_40_R | 2 | AAGCCCGCTAAAGAAGAAGCAC |
| MPXV_Seq_44_F | 2 | CGTAGTAGACGTCTATTCGCCC |
| MPXV_Seq_46_R | 2 | TCCACATGCCAAGTGAAGAGTG |
| MPXV_Seq_50_F | 2 | ACAAGCGAATGAGGGACGAAAA |
| MPXV_Seq_52_R | 2 | CATTTCCATCTCAGACGCGGAA |
| MPXV_Seq_56_F | 2 | TTGCTGTCGAAGAGGCAAAAGA |
| MPXV_Seq_58_R | 2 | CCAACCAGGATGACACGAACAT |
| MPXV_Seq_62_F | 2 | GCAGTCGCGAACAACAACAAAA |
| MPXV_Seq_64_R | 2 | CCGCTCCTCGTTTTTCCCATAA |
| MPXV_Seq_68_F | 2 | TTACATTCCCGTTATTCCGCCC |
| MPXV_Seq_70_R | 2 | AGAGTGTCTCTCGTCCTTGGTT |

Primers are divided into pool 1 (labelled with odd numbers) and pool 2 (labelled with even numbers).

**Table S4.** Primer and probe sequences for pathogen assays used in the ODIN and ODIN-Mpox mobile (Biomeme) qPCR environmental surveillance programs.

| Pathogen Strain | Reference | Oligo Name | Primer/Probe Sequence (5' - 3') |
|---|---|---|---|
| Mpox Congo basin strain (clade I) | Li et al. 2010 | C3L.F | TGTCTACCTGGATACAGAAAGCAA |
| | | C3L.R | GGCATCTCCGTTTAATACATTGAT |
| | | C3L.P | FAM-CCCATATATGCTAAATGTACCGGTACCGGA-3'BHQ1 |
| Mpox Kamituga strain (clade Ib) | Li et al. 2010 | Mpox-K.F | AAGACTTCCAAACTTAATCACTCCT |
| | | Mpox-K.R | CGTTTGATATAGGATGTGGACATTT |
| | | Mpox-K.P | 5'-FAM-ATATTCAGGCGCATATCCACCCACGT- BHQ - 3' |
| Mpox West African strain (clade II) | Li et al. 2010 | G2R_WA.F | CACACCGTCTCTTCCACAGA |
| | | G2R_WA.R | GATACAGGTTAATTTCCACATCG |
| | | G2R_WA.P | FAM-AACCCGTCGTAACCAGCAATACATTT-3'BHQ1 |
| Mpox_Generic (clades I & II) | Li et al. 2010 | G2R_G.F | GGAAAATGTAAAGACAACGAATACAG |
| | | G2R_G.R | GCTATCACATAATCTGGAAGCGTA |
| | | G2R_G.P | FAM-AAGCCGTAATCTATGTTGTCTATCGTGTCC-3'BHQ1 |
| Orthopoxvirus Generic (clades I & II) | Fan et al. 2024 | E9L.F | TCAACTGAAAAGGCCATCTATGA |
| | | E9L.R | GAGTATAGAGCACTATTTCTAAATCCCA |
| | | E9L.P | 5'-FAM-CCATGCAATATACGTACAAGATAGTAGCCAAC- BHQ - 3' |
| *K. pneumoniae* | Hartman et al. 2009 | kheF | GATGAAACGACCTGATTGCATTC |
| | | kheR | CCGGGCTGTCGGGATAAG |
| | | kheP | Cy5-CGCGAACTGGAAGGGCCCG-BHQ2 |
| S. Ttyphi | Nga et al. 2010 | STadhF | CGCGAAGTCAGAGTCGACATAG |
| | | STadhR | AAGACCTCAACGCCGATCAC |
| | | STadhP | FAM-CATTTGTTCTGGAGCAGGCTGACGG-BQ1 |
| Toxigenic *V. cholerae* | Blackstone et al. 2007 | ctxA.F | TTTGTTAGGCACGATGATGGAT |

**Formatted:** Font: Italic

**Formatted:** Font: Italic

| | | | |
|---|---|---|---|
| | | ctxA.R | ACCAGACAATATAGTTTGACCCACTAAG |
| | | ctxA.P | TxRed-TGTTTCCACCTCAATTAGTTTGAGAAGTGCCC-BHQ2 |
| *E. coli* | Silkie et al. 2008 | uidA.F | CGGAAGCAACGCGTAAACTC |
| | | uidA.R | TGAGCGTCGCAGAACATTACA |
| | | uidA.P | Cy5-CGCGTCCGATCACCTGCGTC-BHQ2 |
| Poliovirus | Gerloff et al. 2017 | WPV1F | GTACAAACCAGTCAYGTNAT |
| | | WPV1R | GAGAATAAYTTGTCYTTKGAYGT |
| | | WPV1P | TxRed-CATWATGGTTACRCAMGCACCT-BHQ2 |
| Rotavirus species A | Logan et al. 2006 | RotaA.fwd1 | GGATGTCCTGTACTCCTTGTCAAAA |
| | | RotaA.rev1 | TCCAGTTTGGAACTCATTTCCA |
| | | RotaA.probe1 | Cy5-ATAATGTGCCTTCGACAAT-BHQ2 |
| Hepatitis A | Persson et al. 2021 | havF | CTCTTTGATCTTCCACAAGRGGT |
| | | havR | GCCGCTGTTACCCTATCCAA |
| | | havP | Cy5-AGGCTACGGGTGAAAC-BHQ2 |

**Formatted:** Font: Italic

**Table S5.** Target sequences for pathogen assays in ODIN and ODIN-Mpox mobile (Biomeme) qPCR environmental surveillance.

| Pathogen Strain | Assay | Target Sequence (5' - 3') |
|---|---|---|
| Mpox Congo basin variant (clade I) | C3L | TCTCGAGGCGATGGGCATCTCCGTTTAATACATTGATTAAAGAGTGTCCATCCGGTACCGGTACATTTAGCATATATGGGTCCCATTTTTTGCTTTCTGTATCCAGGTAGACATAGATATTCTATAGTGTCTCC |
| Mpox Kamituga variant (clade Ib) | Mpox-K | AATATTTGAAACACGGCACTTCGAAATGGAAAAGACTTCCAAACTTAATCACTCCTAGATATTCAGGCGCATATCCACCCACGTGTCAGATTGTTAAATGTCCACATCCTATATCAAACGGAAAACTTCTAGCGGCTTAA |
| Mpox West African variant (clade II) | G2R_WA | GGTCCCGGAACATATTCTCACACCGTCTCTTCCACAGATAAATGCGAACCCGTCGTAACCAGCAATACATTTAACTATATCGATGTGGAAATTAACCTGTATCCAGTCAACGACACATCGTGTACTCGG |
| Mpox Generic (clades I & II) | G2R_G | GGCGTACATTGTGTATTAGTCTTGCTATCACATAATCTGGAAGCGTAAGTTCCCGGAGGACACGATAGACAACATAGATTACGGCTTCTGTATTCGTTGTCTTTACATTTTCCATTGGATGGTGC |
| Orthopox Generic (clades I & II) | E9L | GAGTATAGAGCACTATTTCTAAATCCCATCAGACCATATACTGAGTTGGCTACTATCTTGTACGTATATTGCATGGAATCATAGATGGCCTTTTCAGTTGA |
| *K. pneumoniae* | khe | CTCATTTTCGGGAGAAAACGATGAAACGACCTGATTGCATTCGCCACTGGCGCGAACTGGAAGGGCCCGACGATGCCACTTATCCCGACAGCCCGGAGCGTTTTTCGATTGGCGCGCCGCTGGGGCGCGGTT |
| *S.* Typhi | STadh | CGCGAAGTCAGAGTCGACATAGGCATAGATTTTCAGGCCATACATTAATTTGCCAAGGTTGCTATAAACATTTGTTCTGGAGCAGGCTGACGGAAATTCCGTGAACTCGCTGGTGATCGGCGTTGAGGTCTTATCAAACCAGAAGGTGCTTGAGGTCATATTCTCTGCTGGCTGA |
| Toxigenic *V. cholerae* | ctxA | ACCAGACAATATAGTTTGACCCACTAAGTGGGCACTTCTCAAACTAATTGAGGTGGAAACATATCCATCATCGTGCCTAACAAA |
| *E. coli* | uidA | CGGAAGCAACGCGTAAACTCGACCCGACGCGTCCGATCACCTGCGTCAATGTAATGTTCTGCGACGCTCA |
| Poliovirus | WPV1 | GTACAAACCAGGCACGTTATACAGCATAGGACAAGATCAGAGTCCAGTATAGAATCCTTCTTCGCACGCGGTGCTTGTGTAACCATTATGACCGTGGACAACTCGGCATCTACTACGTCCAAGGACAAGTTATTCTC |
| Rotavirus (VP6) | RotaA | GGATGTCCTGTACTCCTTGTCAAAAACTCTTAAAGATGCTAGAGACAAAATTGTCGAAGGCACATTATACTCTAATGTGAGTGATCTAATTCAACAATTTAACCAAATGATAATTACTATGAATGGAAATGAGTTCCAAACTGGA |
| Hepatitis A | hav | CTCTTTGATCTTCCACAAGGGGTAGGCTACGGGTGAAACCCCTTAGGCTAATACTTCTATGAAGAGATGCTTTGGATAGGGTAACAGCGGC |

**Formatted:** Font: Italic

**Formatted:** Font: Italic

**Formatted:** Font: Italic

**Formatted:** Font: Italic

**Table S6.** List of potential pathogens considered in this study.

| No. | Pathogen | Group | Sample Type | Rationality for surveillance |
|---|---|---|---|---|
| 1 | *Klebsiella pneumoniae* | Bacterium | Wastewater | Opportunistic pathogen; hospital-associated; AMR surveillance |
| 2 | *Enterobacter cloacae* | Bacterium | Wastewater | Opportunistic; fecal contamination; AMR |
| 3 | *Streptococcus agalactiae* | Bacterium | Wastewater | Neonatal pathogen; community prevalence via WBE |
| 4 | *Shigella flexneri* | Bacterium | Water/Wastewater | Waterborne dysentery; outbreak detection |
| 5 | *Salmonella enterica* | Bacterium | Water/Wastewater | Water/foodborne outbreaks; WBE |
| 6 | *Coxiella burnetii* | Bacterium | Wastewater | Zoonotic (Q fever); DNA may appear in sewage |
| 7 | *Acinetobacter baumannii* | Bacterium | Wastewater | Hospital wastewater; multidrug resistance |
| 8 | *Escherichia coli* | Bacterium | Water/Wastewater | Primary fecal indicator; pathogenic strains (EHEC, ETEC) |
| 9 | *Neisseria meningitidis* | Bacterium | Wastewater | Respiratory pathogen; community trends via WBE |
| 10 | *Vibrio cholerae* | Bacterium | Water/Wastewater | Cholera surveillance; toxigenic strains |
| 11 | *Shigella dysenteriae* | Bacterium | Water/Wastewater | Severe dysentery; low infectious dose |
| 12 | *Clostridium perfringens* | Bacterium | Water/Soil/Wastewater | Spore-former; persistent fecal pollution marker |
| 13 | *Chlamydia trachomatis* | Bacterium | Wastewater | STI; intracellular but DNA detectable in WBE |
| 14 | *Cryptococcus neoformans* | Fungus | Soil/Wastewater | Environmental yeast; bird-dropping/soil; opportunistic |
| 15 | *Pseudomonas aeruginosa* | Bacterium | Water/Wastewater | Opportunistic; biofilms in water systems; AMR |
| 16 | *Enterococcus faecium* | Bacterium | Water/Wastewater | Fecal indicator; VRE/AMR surveillance |
| 17 | *Francisella tularensis* | Bacterium | Wastewater | Zoonotic; rare but detectable by DNA |
| 18 | Streptococcus pneumoniae | Bacterium | Wastewater | Respiratory pathogen; WBE |
| 19 | Staphylococcus epidermidis | Bacterium | Wastewater | Opportunistic skin flora; device infections; AMR |
| 20 | *Streptococcus pyogenes* | Bacterium | Wastewater | Group A strep; community trends via WBE |
| 21 | *Rhizopus microsporus* | Fungus | Soil | Environmental mold; rare human infections |
| 22 | *Salmonella bongori* | Bacterium | Wastewater | Less common Salmonella; fecal pollution |
| 23 | *Vibrio mimicus* | Bacterium | Water/Wastewater | Estuarine pathogen; gastroenteritis |
| 24 | *Neisseria gonorrhoeae* | Bacterium | Wastewater | STI; trends via wastewater genomics |
| 25 | *Serratia marcescens* | Bacterium | Wastewater | Opportunistic; nosocomial; AMR |
| 26 | *Legionella pneumophila* | Bacterium | Drinking Water | Premise plumbing; aerosol transmission |

| | | | | |
|---|---|---|---|---|
| 27 | *Mucor indicus* | Fungus | Soil | Environmental mold; rare mucormycosis |
| 28 | *Leptospira interrogans* | Bacterium | Water/Soil | Leptospirosis; rodent-urine contamination |
| 29 | *Candida albicans* | Fungus | Wastewater | Opportunistic yeast; AMR monitoring |
| 30 | *Clostridioides difficile* | Bacterium | Wastewater | Healthcare-associated; spores; AMR |
| 31 | *Staphylococcus aureus* | Bacterium | Wastewater | MRSA surveillance; skin/nasal shedding |
| 32 | *Francisella hispaniensis* | Bacterium | Wastewater | Rare Francisella; environmental DNA |
| 33 | *Campylobacter jejuni* | Bacterium | Water/Wastewater | Leading bacterial gastroenteritis; waterborne |
| 34 | *Enterococcus faecalis* | Bacterium | Water/Wastewater | Fecal indicator; AMR |
| 35 | *Vibrio vulnificus* | Bacterium | Water/Wastewater | Severe wound/septicemia; warm brackish waters |
| 36 | *Proteus mirabilis* | Bacterium | Wastewater | Opportunistic; AMR surveillance |
| 37 | *Vibrio alginolyticus* | Bacterium | Water/Wastewater | Marine pathogen; wound/ear infections |
| 38 | *Helicobacter pylori* | Bacterium | Wastewater | Fecal shedding suspected; WBE |
| 39 | *Bacillus anthracis* | Bacterium | Soil | Spore-former; soil persistence; anthrax risk |
| 40 | *Vibrio parahaemolyticus* | Bacterium | Water/Wastewater | Seafood-borne gastroenteritis |
| 41 | *Trichophyton rubrum* | Fungus | Soil | Dermatophyte; environmental presence |
| 42 | *Vibrio cincinnatiensis* | Bacterium | Water/Wastewater | Rare Vibrio; environmental presence |
| 43 | *Vibrio fluvialis* | Bacterium | Water/Wastewater | Waterborne diarrhea; estuarine waters |
| 44 | *Chlamydia pneumoniae* | Bacterium | Wastewater | Respiratory chlamydia; WBE |
| 45 | *Vibrio damsela* (Photobacterium damselae subsp. damselae) | Bacterium | Water/Wastewater | Marine pathogen; wound infections |
| 46 | *Vibrio hollisae (Grimontia hollisae)* | Bacterium | Water/Wastewater | Marine; gastroenteritis |
| 47 | *Mucor circinelloides* | Fungus | Soil | Environmental mold; mucormycosis |
| 48 | Mycobacterium tuberculosis | Bacterium | Wastewater | TB DNA in sewage; WBE |
| 49 | *Cryptosporidium* | Protozoan | Water/Wastewater | Waterborne oocysts; resistant to chlorine |
| 50 | *Gardia* | Protozoan | Water/Wastewater | (Giardia) classic waterborne; low infectious dose |
| 51 | *Vibrio furnissii* | Bacterium | Water/Wastewater | Marine; gastroenteritis |
| 52 | *Vibrio metschnikovii* | Bacterium | Water/Wastewater | Rare; aquatic environments |
| 53 | *Vibrio harveyi* | Bacterium | Water/Wastewater | Marine; opportunistic human cases |
| 54 | *Vibrio navarrensis* | Bacterium | Water/Wastewater | Environmental Vibrio; occasional infections |

| | | | | |
|---|---|---|---|---|
| 55 | *Mycobacterium leprae* | Bacterium | Wastewater | Leprosy DNA in sewage; WBE |
| 56 | *Haemophilus influenzae* | Bacterium | Wastewater | Respiratory pathogen; WBE |
| 57 | *Rickettsia rickettsii* | Bacterium | Wastewater | Tick-borne; rare DNA in sewage |
| 58 | *Borrelia burgdorferi* | Bacterium | Wastewater | Lyme agent; rare DNA in sewage |
| 59 | *Treponema pallidum* | Bacterium | Wastewater | Syphilis; fragile but DNA detectable |
| 60 | *Listeria monocytogenes* | Bacterium | Water/Soil | Food safety; environmental persistence |
| 61 | *Clostridium tetani* | Bacterium | Soil | Tetanus spores; soil persistence |
| 62 | *Clostridium botulinum* | Bacterium | Soil/Water | Botulinum spores/toxins; environmental risk |
| 63 | *Yersinia pestis* | Bacterium | Soil/Wastewater | Plague agent; rare environmental DNA |
| 64 | *Brucella melitensis* | Bacterium | Wastewater | Zoonotic; occupational settings; DNA |
| 65 | *Candida auris* | Fungus | Wastewater | Emerging MDR yeast; outbreak tracking |
| 66 | *Candida glabrata* | Fungus | Wastewater | Opportunistic; AMR |
| 67 | *Cryptococcus gattii* | Fungus | Soil | Environmental; causes cryptococcosis |
| 68 | *Aspergillus fumigatus* | Fungus | Soil | Spores; respiratory disease; azole resistance |
| 69 | *Aspergillus flavus* | Fungus | Soil | Aflatoxin producer; opportunistic infections |
| 70 | *Histoplasma capsulatum* | Fungus | Soil | Guano-enriched soils; endemic mycosis |
| 71 | *Blastomyces dermatitidis* | Fungus | Soil | Endemic mycosis; soil detection |
| 72 | *Coccidioides immitis* | Fungus | Soil | Valley fever; arid soils |
| 73 | *Coccidioides posadasii* | Fungus | Soil | Valley fever; arid soils |
| 74 | *Pneumocystis jirovecii* | Fungus | Wastewater | Opportunistic; DNA in sewage |
| 75 | *Trichophyton mentagrophytes* | Fungus | Soil | Dermatophyte; environmental presence |
| 76 | *Microsporum canis* | Fungus | Soil | Dermatophyte; zoonotic |
| 77 | *Sporothrix schenckii* | Fungus | Soil | Sporotrichosis; soil/plant material |
| 78 | *Plasmodium falciparum* | Protozoan | Wastewater | Vector-borne but DNA detectable for WBE |
| 79 | *Plasmodium vivax* | Protozoan | Wastewater | WBE for malaria burden |
| 80 | *Plasmodium malariae* | Protozoan | Wastewater | WBE for malaria burden |
| 81 | *Plasmodium ovale* | Protozoan | Wastewater | WBE for malaria burden |
| 82 | *Plasmodium knowlesi* | Protozoan | Wastewater | Zoonotic malaria; DNA in sewage |

| | | | | |
|---|---|---|---|---|
| 83 | *Toxoplasma gondii* | Protozoan | Soil/Wastewater | Oocysts in environment; waterborne risk |
| 84 | *Entamoeba histolytica* | Protozoan | Water/Wastewater | Waterborne amoebiasis |
| 85 | *Cryptosporidium parvum* | Protozoan | Water/Wastewater | Chlorine-resistant oocysts; outbreaks |
| 86 | *Cyclospora cayetanensis* | Protozoan | Water/Wastewater | Produce/water outbreaks |
| 87 | *Trichomonas vaginalis* | Protozoan | Wastewater | STI; DNA detection for WBE |
| 88 | *Trypanosoma brucei* | Protozoan | Wastewater | Vector-borne; DNA detectable |
| 89 | *Trypanosoma cruzi* | Protozoan | Wastewater | Chagas; DNA detectable |
| 90 | *Leishmania donovani* | Protozoan | Wastewater | Visceral leishmaniasis; DNA detectable |
| 91 | *Leishmania major* | Protozoan | Wastewater | Cutaneous leishmaniasis; DNA detectable |
| 92 | *Ascaris lumbricoides* | Helminth | Soil/Wastewater | Soil-transmitted helminth; sanitation indicator |
| 93 | *Trichuris trichiura* | Helminth | Soil/Wastewater | Soil-transmitted helminth |
| 94 | *Ancylostoma duodenale* | Helminth | Soil/Wastewater | Hookworm; soil transmission |
| 95 | *Necator americanus* | Helminth | Soil/Wastewater | Hookworm; soil transmission |
| 96 | *Strongyloides stercoralis* | Helminth | Soil/Wastewater | Soil-transmitted helminth |
| 97 | *Taenia saginata* | Helminth | Wastewater | Eggs in feces; sanitation monitoring |
| 98 | *Taenia solium* | Helminth | Wastewater | Neurocysticercosis risk; sanitation |
| 99 | *Echinococcus granulosus* | Helminth | Wastewater | Eggs in feces; zoonotic |
| 100 | *Echinococcus multilocularis* | Helminth | Wastewater | Eggs in feces; zoonotic |
| 101 | *Schistosoma mansoni* | Helminth | Water/Wastewater | Snail-transmitted; endemic areas |
| 102 | *Schistosoma haematobium* | Helminth | Water/Wastewater | Urinary schistosomiasis; endemic |
| 103 | *Schistosoma japonicum* | Helminth | Water/Wastewater | Endemic regions; zoonotic reservoirs |
| 104 | *Enterobius vermicularis* | Helminth | Wastewater | Eggs in feces; childcare settings |
| 105 | *Salmonella Ttyphi* | Bacterium | Water/Wastewater | Water/foodborne outbreaks; WBE |

**Table S7.** Concentration and purity of nucleic acids extracted from wastewater using ZymoBIOMICS DNA Miniprep Kit and ML NA extraction method.

| Sample ID | Sample | Extraction Kit | Qubit (ng/µL) | Nanodrop (ng/µL) | 260/280 | 260/230 |
|---|---|---|---|---|---|---|
| 1 | Wastewater | ZymoBIOMICS | 12.1 | 20.6 | 1.94 | 1.13 |
| 2 | Wastewater | ZymoBIOMICS | 13.3 | 13.6 | 2.15 | 0.66 |
| 3 | Wastewater | ZymoBIOMICS | 13.7 | 15.9 | 2.19 | 0.95 |
| 4 | Wastewater | ZymoBIOMICS | 17.7 | 19.5 | 2.10 | 1.01 |
| 5 | Wastewater | ZymoBIOMICS | 13.9 | 22.9 | 2.04 | 0.89 |
| 7 | Wastewater | ML NA | 22.6 | 32.7 | 2.17 | 2.34 |
| 8 | Wastewater | ML NA | 23.3 | 43.5 | 2.19 | 2.43 |
| 9 | Wastewater | ML NA | 14.0 | 27.7 | 2.21 | 2.40 |
| 10 | Wastewater | ML NA | 19.5 | 28.4 | 2.18 | 2.31 |
| 11 | Wastewater | ML NA | 22.4 | 38.2 | 2.18 | 2.29 |

Table S8 Pilot testing will use field samples from Tanzania categorized as qPCR-positive (1) or negative (0). A dash (—) denotes data not determined.

| Target/Sample ID | M1 | M2 | M3 | M4 | M5 | M6 | M7 | M8 | M9 | M10 | M11 | M12 | M13 | M14 | M15 |
|---|---|---|---|---|---|---|---|---|---|---|---|---|---|---|---|
| Target/ Sample Type | DW | DW | DW | WW_T | WW_T | WW_T | WW_S | WW_S | WW_S | WW_U | WW_U | WW_U | Soil | Soil | Soil |
| *K. pneumoniae* (khe) | 0 | 1 | 0 | 0 | 1 | 1 | 1 | 1 | 1 | 1 | 0 | 0 | 1 | 0 | 0 |
| Salmonella Ttyphi (STadh) | — | 0 | — | — | — | 0 | — | — | 0 | 0 | — | — | 0 | — | — |
| *Vibrio cholerae* (ctxA) | 0 | 0 | 0 | 0 | 0 | 0 | 0 | 0 | 0 | 1 | 0 | 0 | 0 | 1 | 0 |
| *Escherichia coli* (uidA) | — | — | — | — | — | — | — | — | — | — | — | — | — | — | — |
| Wild pPoliovirus-1 (WPV1) | 0 | — | — | — | 0 | 0 | 0 | 0 | 0 | 0 | 0 | 0 | 0 | 0 | — |
| Rotavirus Spp A (rotaA) | — | — | 1 | 1 | 1 | 1 | 0 | 0 | 0 | 0 | 0 | 0 | 0 | 0 | 0 |
| Hepatitis A virus (hav) | — | — | — | — | — | 1 | — | — | 1 | 0 | — | — | 0 | — | — |
| Mpox clade II WA (G2R_WA) | — | — | — | — | — | — | — | — | — | 0 | 0 | 0 | — | — | — |
| Mpox clade I Congo Basin (C3L) | — | — | — | — | — | — | — | — | — | — | — | — | — | — | — |
| Mpox clade Ib (Mpox-K) | 0 | — | — | — | — | 0 | — | — | 0 | 0 | — | — | 0 | — | — |
| Generic Mpox clade I & II (G2R_G) | 0 | — | — | — | — | 0 | — | — | 0 | 0 | — | — | 0 | — | — |
| Generic Orthopox clade I & II (E9L) | — | — | — | — | — | — | — | — | — | — | — | — | — | — | — |

**Formatted:** Font: Italic

**Formatted:** Font: Italic

**Formatted:** Font: Italic
